# Role of endogenous adenine in kidney failure and mortality with diabetes

**DOI:** 10.1101/2023.05.31.23290681

**Authors:** Kumar Sharma, Guanshi Zhang, Jens Hansen, Petter Bjornstad, Hak Joo Lee, Rajasree Menon, Leila Hejazi, Jian-Jun Liu, Anthony Franzone, Helen C. Looker, Byeong Yeob Choi, Roman Fernandez, Manjeri A. Venkatachalam, Luxcia Kugathasan, Vikas S. Sridhar, Loki Natarajan, Jing Zhang, Varun Sharma, Brian Kwan, Sushrut Waikar, Jonathan Himmelfarb, Katherine Tuttle, Bryan Kestenbaum, Tobias Fuhrer, Harold Feldman, Ian H. de Boer, Fabio C. Tucci, John Sedor, Hiddo Lambers Heerspink, Jennifer Schaub, Edgar Otto, Jeffrey B. Hodgin, Matthias Kretzler, Christopher Anderton, Theodore Alexandrov, David Cherney, Su Chi Lim, Robert G. Nelson, Jonathan Gelfond, Ravi Iyengar, the Kidney Precision Medicine Project

## Abstract

Diabetic kidney disease (DKD) can lead to end-stage kidney disease (ESKD) and mortality, however, few mechanistic biomarkers are available for high risk patients, especially those without macroalbuminuria. Urine from participants with diabetes from Chronic Renal Insufficiency Cohort (CRIC), Singapore Study of Macro-Angiopathy and Reactivity in Type 2 Diabetes (SMART2D), and the Pima Indian Study determined if urine adenine/creatinine ratio (UAdCR) could be a mechanistic biomarker for ESKD. ESKD and mortality were associated with the highest UAdCR tertile in CRIC (HR 1.57, 1.18, 2.10) and SMART2D (HR 1.77, 1.00, 3.12). ESKD was associated with the highest UAdCR tertile in patients without macroalbuminuria in CRIC (HR 2.36, 1.26, 4.39), SMART2D (HR 2.39, 1.08, 5.29), and Pima Indian study (HR 4.57, CI 1.37-13.34). Empagliflozin lowered UAdCR in non-macroalbuminuric participants. Spatial metabolomics localized adenine to kidney pathology and transcriptomics identified ribonucleoprotein biogenesis as a top pathway in proximal tubules of patients without macroalbuminuria, implicating mammalian target of rapamycin (mTOR). Adenine stimulated matrix in tubular cells via mTOR and stimulated mTOR in mouse kidneys. A specific inhibitor of adenine production was found to reduce kidney hypertrophy and kidney injury in diabetic mice. We propose that endogenous adenine may be a causative factor in DKD.

## Introduction

Progression to organ failure is marked by fibrosis and loss of architecture in solid organs, such as the kidney. In almost all progressive chronic kidney diseases (CKD) the features that are most consistently associated with functional loss of the glomerular filtration rate (GFR) are the degree of glomerulosclerosis, tubulointerstial fibrosis, vascular injury, and proteinuria (1-4). However, many patients who eventually develop end-stage kidney disease (ESKD) are non-proteinuric at the time impaired GFR is recognized. Non-proteinuria is defined as urine albumin-to-creatinine ratio (ACR) ≤ 300 mg/creatinine or urine albumin excretion ≤ 300mg/day (5). As non-proteinuric or non-macroalbuminuric DKD accounts for >40% of prevalent ESKD in patients with type 2 diabetes (5-7) and 75% of prevalent CKD (GFR <60 mL/min/1.73m^2^) (8) identifying the patients at risk for progression in early stages of disease is an important step to improve clinical outcomes. This is especially relevant as the armamentarium of therapies for DKD to mitigate kidney disease progression has rapidly expanded (9-11).

Establishing novel biomarkers that predict progression and represent biologically relevant pathways in DKD could improve the care of patients with diabetes. To identify novel biomarkers, we recently performed an untargeted urine metabolomics study in patients with type 2 diabetes (T2D) and impaired eGFR from the Chronic Renal Insufficiency Cohort (CRIC) study (12) and identified 15 candidate metabolites associated with ESKD. A targeted assay validated 13 of these metabolites, one of which was adenine. As exogenous adenine has been found to cause kidney failure in mice, rats, and dogs (13-15), we evaluated whether endogenous adenine could play a role in progression of kidney disease in patients with diabetes.

## Results

### Urine adenine/creatinine ratio predicts kidney failure and all cause mortality in the CRIC and SMART2D cohorts

The baseline clinical characteristics of the participants with diabetes from CRIC and SMART2D are shown in Table 1. Of the 904 subjects evaluated from CRIC, 558 had either normoalbuminuria or microalbuminuria, 341 had macroalbuminuria, and 5 had no data for 24h albumin. The mean eGFR was 40 mL/min/1.73m^2^. The top tertile of baseline urine adenine/creatinine ratio (UAdCR) was found to identify the participants with diabetes who were at high risk for ESKD and all cause mortality (adjusted HR 1.57, 95% CI 1.18, 2.10, as compared to the lowest tertile) (Figure 1A) and a similar significant relationship was found using UAdCR as a continuous variable (Table 2). The value of the top tertile of UAdCR to identify patients with diabetes at high risk for ESKD and all cause mortality was confirmed in participants from SMART2D who had reduced eGFR and normoalbuminuria or microalbuminuria (adjusted HR 1.77, 95% CI 1.00, 3.12) (Figure 1B, Table 2, datasets combined in Supplementary Figure 2A).

**Figure 1.**
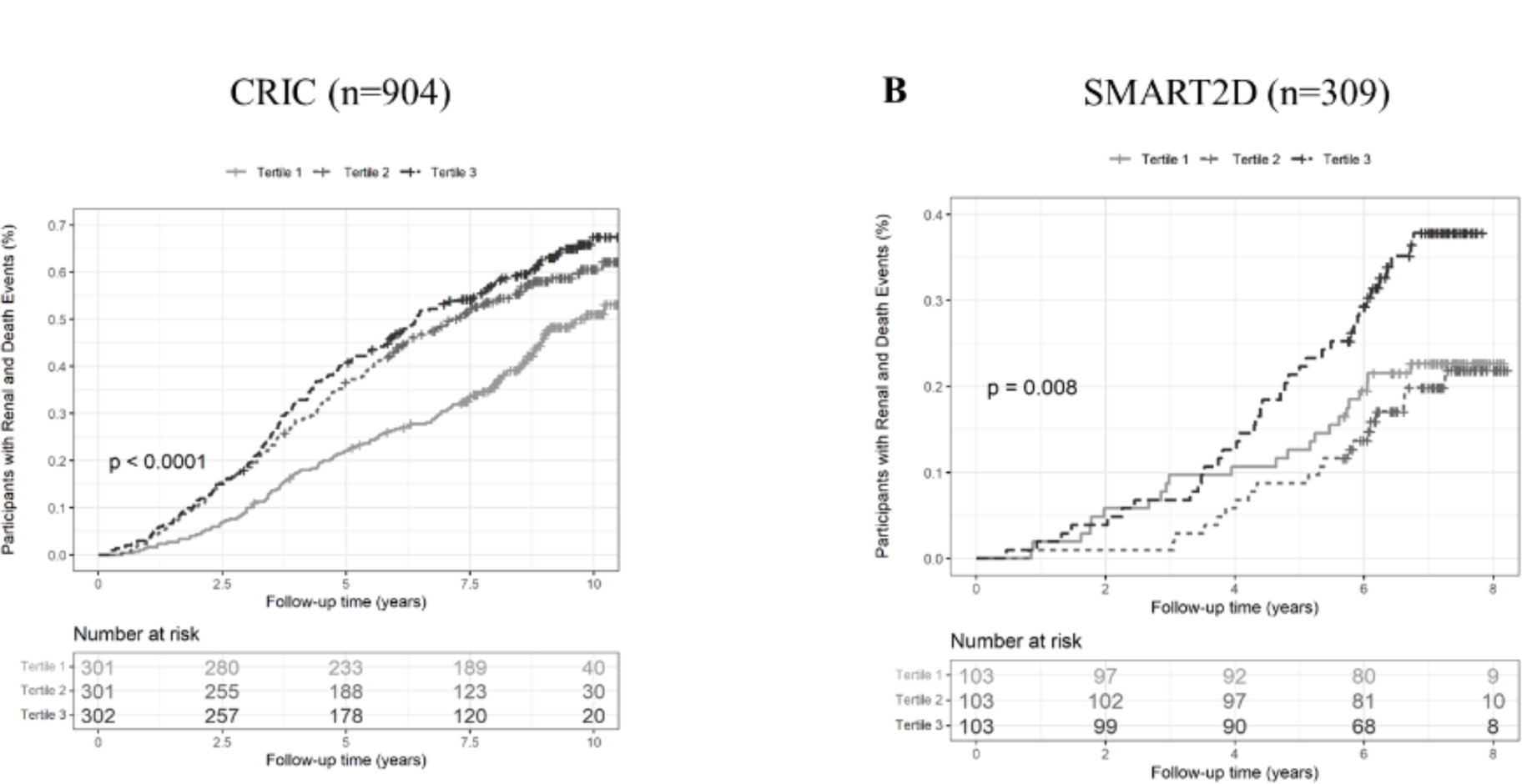
High urine adenine/creatinine (UAdCR) levels identify patients with diabetes who are at high risk of end stage kidney disease (ESKD) and mortality. Participants with diabetes in the Chronic Renal Insufficiency Cohort (CRIC) cohort (n=904) had UAdCR measured within 1 year of enrollment and followed for 10 years. The participants in the top tertile had the highest risk of ESKD and all cause mortality (A). Participants from the Singapore Study of Macro-Angiopathy and microvascular Reactivity in Type 2 Diabetes (SMART2D) study (n=309) had UAdCR measurements at the time of enrollment and were followed for 7 years. The participants in the top tertile for UAdCR had the highest risk for ESKD and all-cause mortality.

**Table 1.**
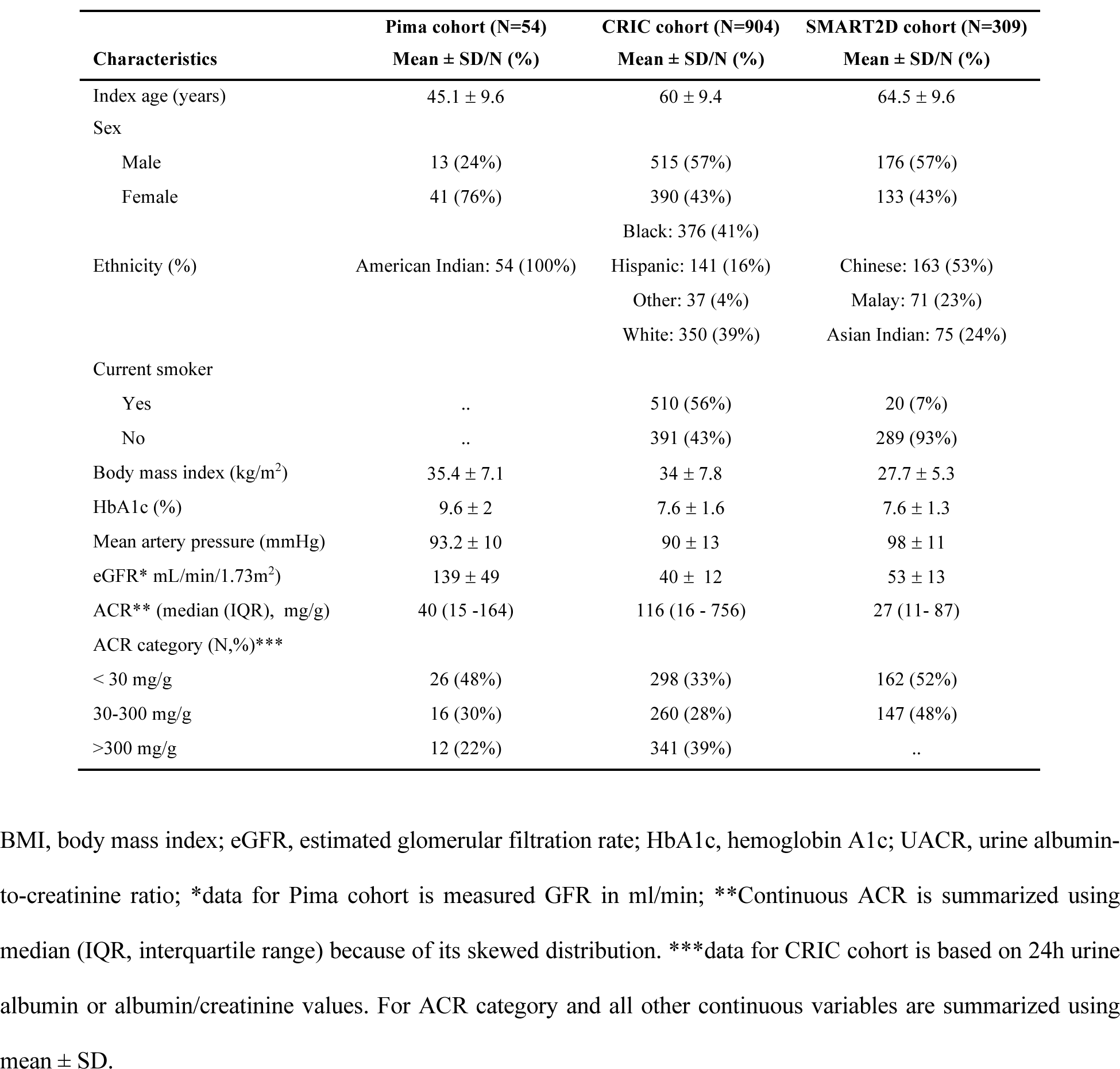
Baseline characteristics of patients with diabetes in the Pima American Indians, CRIC and SMART2D studies.

**Table 2.** Association of baseline urine adenine/creatinine ratio (UAdCR) with risk for progression to ESKD and all-cause mortality in CRIC and SMART2D participants with type 2 diabetes with 7 years follow up.

| Adenine/Creatinine ratio | CRIC cohort (N=889) |  | SMART2D cohort (N=309) |  |
| --- | --- | --- | --- | --- |
|  | HR (95% CI) | p value | HR (95% CI) | p value |
| 1-SD increment | 1.15 (1.03-1.28) | 0.010 | 1.48 (1.15-1.90) | 0.003 |
| Tertile 2 vs tertile 1 | 1.59 (1.21-2.09) | <0.001 | 0.81 (0.44-1.50) | 0.502 |
| Tertile 3 vs tertile 1 | 1.57 (1.18-2.10) | 0.002 | 1.77 (1.00-3.12) | 0.048 |

Multivariate Cox proportional hazard regression models were adjusted for baseline age, sex, ethnicity, body mass index, mean arterial pressure, hemoglobin A1c, eGFR and natural-log transformed urine albumin/creatinine ratio. UAdCR was modelled as both continuous variable (1-SD increment in log2-transformed adenine/creatinine ratio) and categorical variable (low tertile as reference). There were 15 subjects in CRIC with missing values for the clinical co-variates.

### UAdCR predicts kidney failure in the non-macroalbuminuric Pima, CRIC, and SMART2D cohorts and empagliflozin reduces UAdCR

The UAdCR was also evaluated in early-stage disease (measured GFR>90 mL/min/1.73m^2^) in a Pima Indian cohort with >20 year follow up (Table 1). As the majority of the participants in the Pima Indian cohort had non-macroalbuminuria (n=42 of the 54 participants), the association of UAdCR with longitudinal progression to ESKD is presented in this non-macroalbuminuric cohort. ESKD was associated with the top UAdCR tertile (HR 4.57, CI 1.37-13.34) (Supplementary Table 1). UAdCR was also measured from 2 untimed spot urine samples obtained 1 year apart and found to be consistent across the individual paired samples (r=0.665, p<0.0001) (Supplementary Figure 3). Similar relationships to predict ESKD was found in the non-macroalbuminuric participants in CRIC (adjusted HR 2.36, 95% CI 1.26, 4.39) and SMART2D (adjusted HR 2.39, 95% CI 1.08, 5.29) (Figure 2A, 2B, Supplementary Table 2, combined datasets in Supplementary Figure 2B). Of note, there were no significant correlations of UAdCR with the UACR or eGFR in the non-macroalbuminuric subjects from CRIC or SMART2D (Supplementary Table 3). Of the CRIC subjects with macroalbuminuria, there were modest associations between the top tertile of UAdCR and ESKD (HR 1.10, CI 0.75, 1.60) and mortality (HR 1.33, CI 0.59, 3.01).

**Figure 2.**
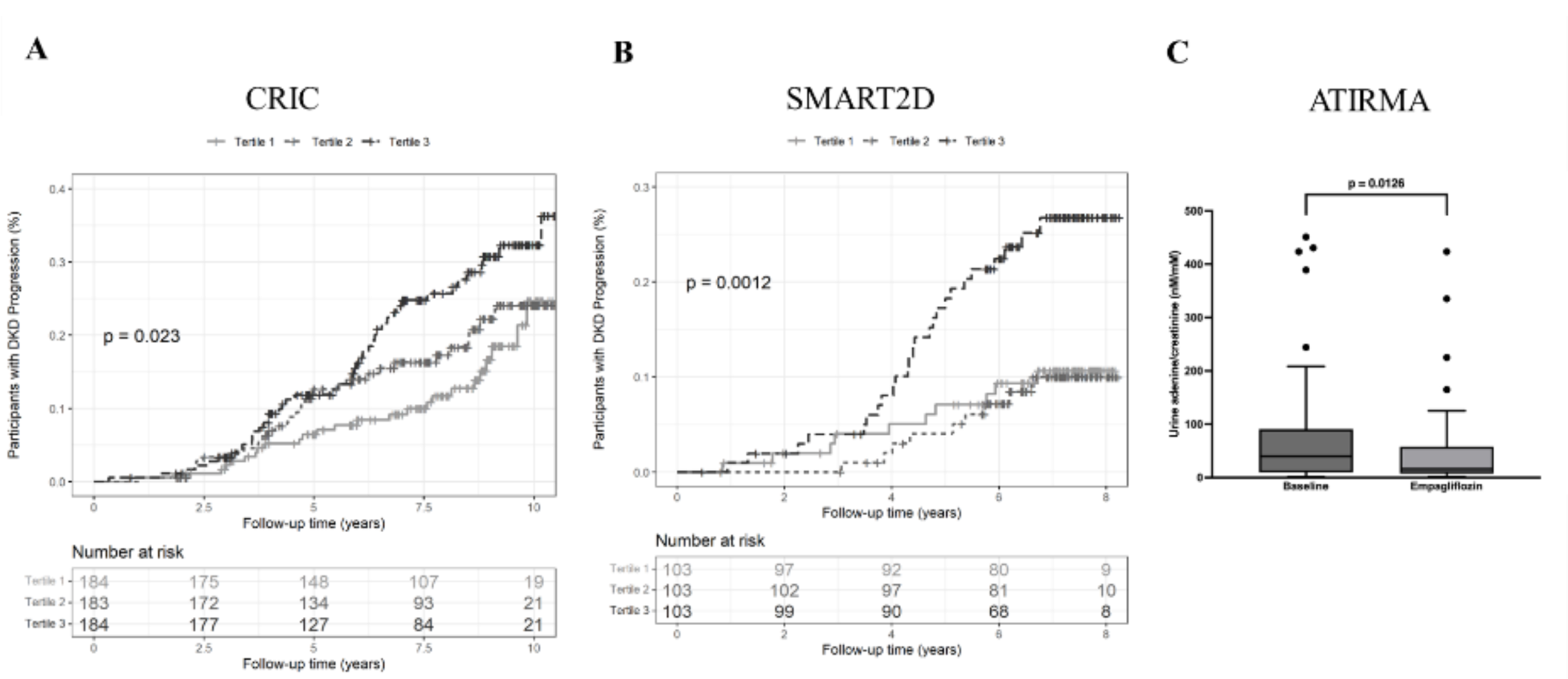
High urine adenine/creatinine (UAdCR) tertile identifies end stage kidney disease (ESKD) outcome in non-macroalbuminuric patients with diabetes and empagliflozin reduced urine adenine/creatinine ratio. The participants with the top UAdCR tertile had a significant increase in risk of ESKD from CRIC (n=551) (A) and SMART2D (n=309) (B) studies. Patients with T1 diabetes underwent treatment with empagliflozin for 8 weeks which reduced UAdCR levels (n=40 patients) (C).

To determine if UAdCR could be modified in non-macroalbuminuric participants with normal or elevated measured GFR by glycemia or a therapeutic intervention with an SGLT2 inhibitor, the UAdCR was measured during euglycemia or hyperglycemia before and after empagliflozin in patients with T1D (clinical characteristics described in Supplementary Table 4). Acute hyperglycemia did not alter UAdCR levels (Supplementary Figure 4), however empagliflozin significantly lowered UAdCR by 36.4% (Figure 2C).

### Adenine is localized to regions of kidney fibrosis and is increased in patients with diabetes

A spatial metabolomics platform was developed to annotate small molecules (<700 Da) and performed on kidney biopsies from healthy controls and in patients with diabetes (clinical characteristics in Supplementary Table 4). Adenine was present at low intensity in normal glomeruli and blood vessels in the healthy control kidney (Figure 3A) and enhanced in regions of arteriolosclerosis, tubulointerstial fibrosis and early glomerulosclerosis in the diabetic kidney (Figure 3B). There was an overall increase in adenine in the whole section of kidney biopsies from participants with diabetes as compared to healthy controls (Figure 3C). The spatial adenine values in rat kidney sections were found to correlate well with the UAdCR in a ZDF diabetic model (r=0.73, *p*<0.001, Supplementary Table 5).

**Figure 3.**
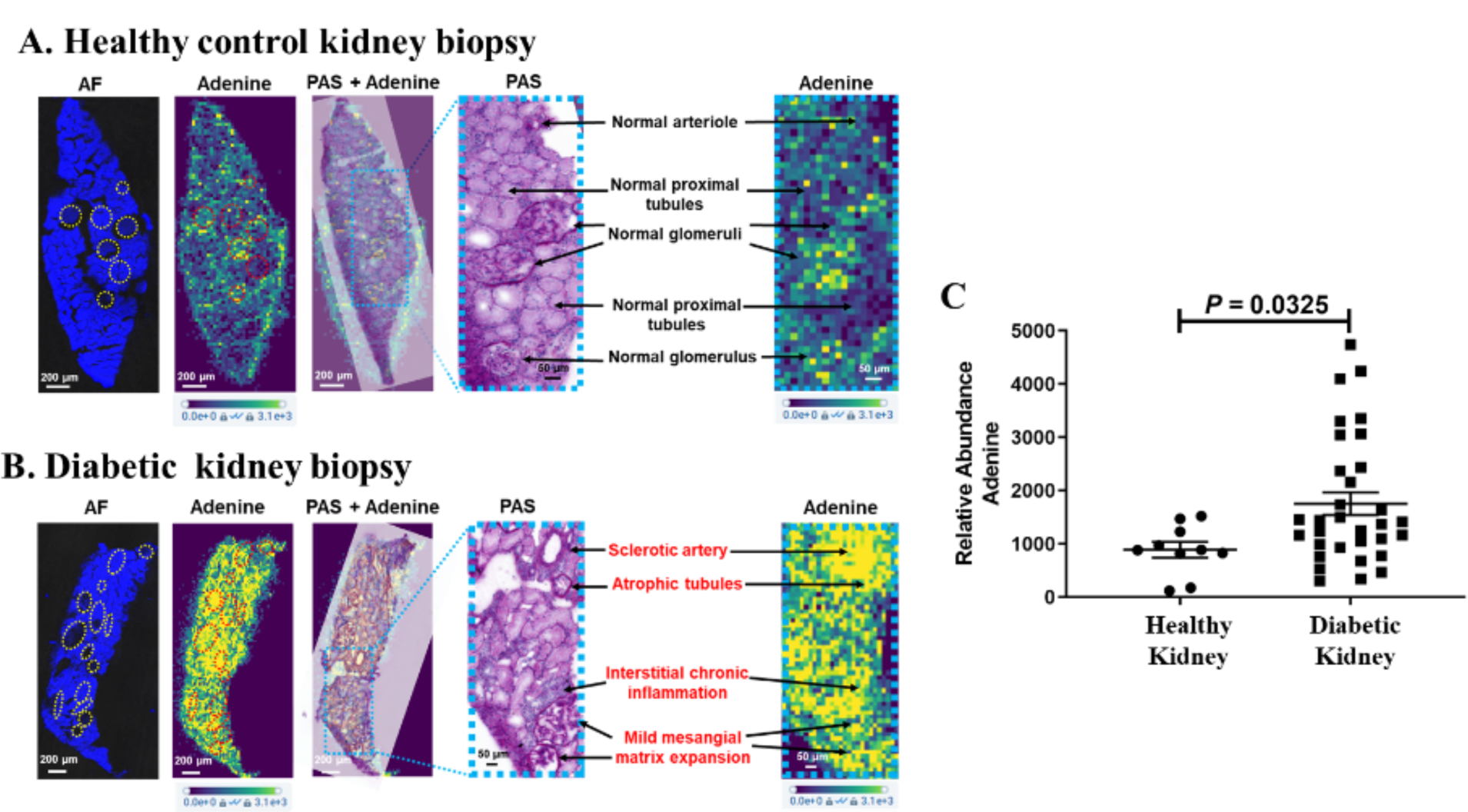
Spatial metabolomics identifies adenine in regions of pathology in non-macroalbuminuric patients with diabetes. Adenine was localized to regions of normal glomeruli and vessels in the normal kidney (A). In a diabetic kidney, adenine is diffusely increased across the tissue section and prominent in regions of sclerotic blood vessels, glomeruli with mild sclerosis and regions of atrophic tubules and interstitial inflammation (B). Quantitative assessment across healthy controls (n=5 from Control of Renal Oxygen Consumption, Mitochondrial Dysfunction and Insulin Resistance (CROCODILE) study) and diabetic samples (n=8 T1D from CROCODILE and n=8 T2D (2 from CROCODILE and 6 from Kidney Precision Medicine Project (KPMP)) demonstrates a statistically significant increase of adenine in kidney tissue sections (C).

### Single cell transcriptomics identify ribonucleoprotein biogenesis as a dominant pathway in non-macroalbuminuric DKD

As adenine was prominent in regions of tubular pathology in the diabetic kidney and empagliflozin treatment lowered the UAdCR in patients, the proximal tubular cells were considered to be a target cell type affected by adenine. Single cell transcriptomics from proximal tubular cells were studied in DKD patients from the KPMP study (n=28) and an unbiased pathway analysis was performed based on differentially regulated genes. The top pathway identified was the ribosomal nucleoprotein biogenesis pathway in patients without macroalbuminuria and low eGFR (Figure 4A, B). In addition, small and large ribosomal subunit organization pathways were also upregulated in these patients. Replication of these results from KPMP was found in the CROCODILE study in diabetic patients without macroabuminuria and normal GFR (Figure 4C). As ribonucleoprotein biogenesis, and small and large ribosomal subunit organization is closely linked to activity of mammalian target of rapamycin (mTOR) (16) and adenine has been found to stimulate mTOR (17), this pathway was evaluated to mediate adenine-induced effects on proximal tubular cells.

**Figure 4.**
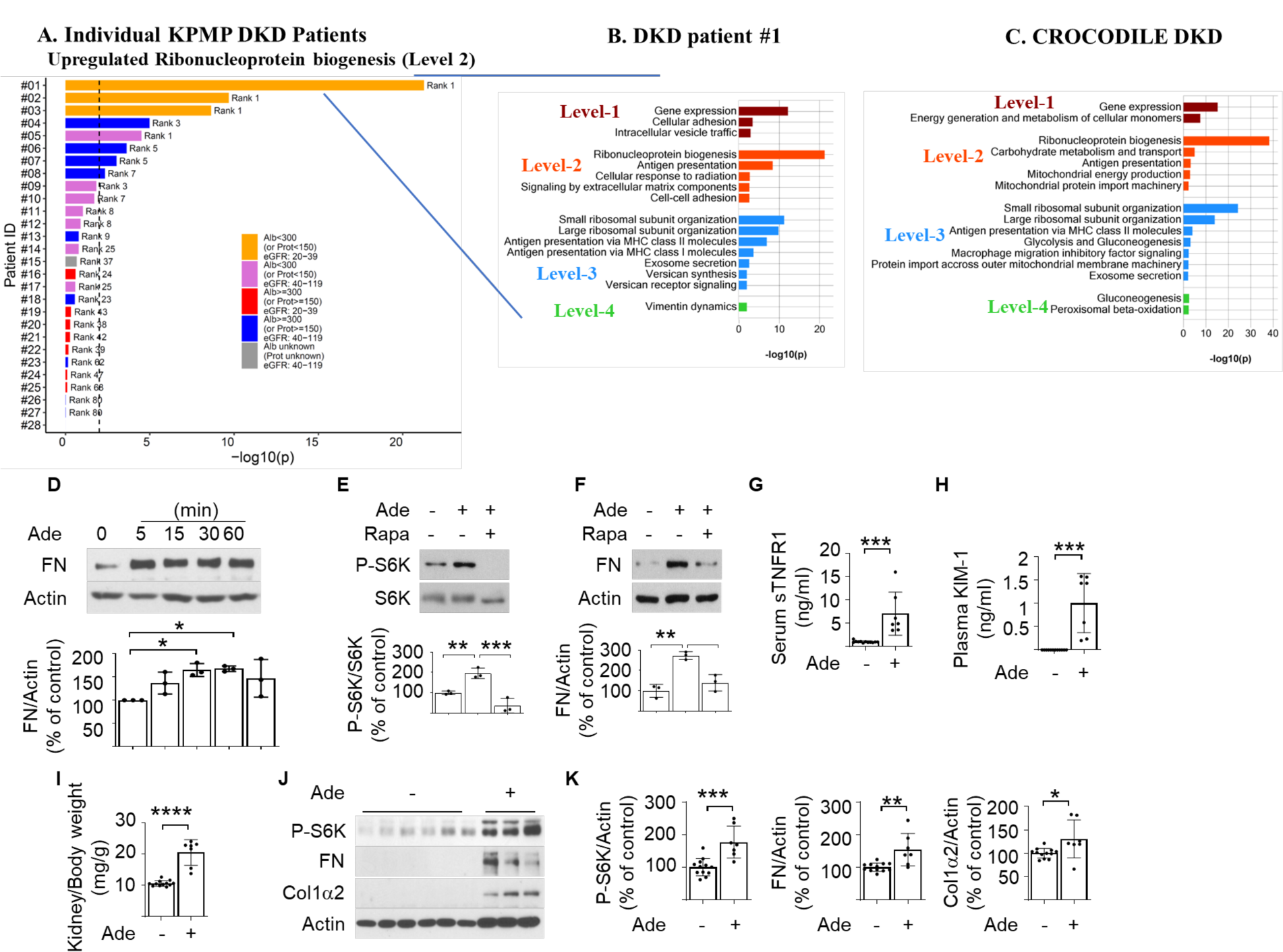
Molecular pathways and events implicating ribonucleoprotein biogenesis and mammalian target of rapamycin (mTOR) pathway with adenine in diabetic kidney disease (DKD). Protein synthesis (Ribonucleoprotein (RNP) biogenesis) pathway increased in proximal tubule cells of DKD patients without proteinuria (A) Single cell-transcriptomic data obtained from DKD kidney biopsies for the Kidney Precision Medicine Project (KPMP) was analyzed for differentially expressed genes in proximal tubule (PT) of each DKD patient versus healthy reference tissue. Upregulated genes with an adjusted p-value ≤ 0.01 and ranked among the top 600 significant DEGs were subjected to pathway enrichment analysis using the Molecular Biology of the Cell Ontology (MBCO). Ranking for the RNP biogenesis pathway (a Level 2 pathway canonically regulated by the mTOR pathway) is shown for 28 individual patients. Vertical dashed line indicates p value ≤ 0.01 for pathway ranking (B) Up to top five, five, ten and five level-1 (dark red), level-2 (red), level-3 (blue) and level-4 (green) pathways using MBCO are shown for patient #1 (p-value ≤ 0.01). See blue lines from A to B. Single cell transcriptomic data from T2D patients (N=6) with low albuminuria compared to cohort specific healthy samples was analyzed to identify upregulated pathways in PT cells (C). Note that the RNP biogenesis pathway is the top ranked Level-2 pathway in both independent studies. Cell culture studies in mouse proximal tubular cells demonstrated an increase in fibronectin (D), phospho-S6 kinase and mediation of fibronectin (FN) upregulation is blocked by rapamycin (E, F), indicating mTOR mediates adenine effect. Adenine administration to mice increases serum soluble tumor necrosis factor-1 (sTNFR1) (G), plasma kidney injury marker-1 (KIM-1) (H), stimulates kidney (I) and matrix molecules in the kidney (J, K) (n=12 in control group and n=7 in adenine treated group, *p<0.05, **p<0.01, ***p<0.001, ****p<0.0001).

### Mechanism of adenine induced matrix production is via the mTOR pathway and adenine increases KIM1 and sTNFR1 in mice

To determine whether adenine could be in the causative pathway for tissue fibrosis, adenine was added to mouse and human proximal tubular cells. There was a robust and early stimulation of fibronectin by adenine (Figure 4D, Supplementary Figure 5A). In addition, adenine stimulated mTOR activity as demonstrated by enhanced phosphorylation of S6 kinase (Figure 4E, Supplementary Figure 5B). Inhibition of mTORC1 with rapamycin blocked adenine-induced production of fibronectin (Figure 4E, 4F). Exposure of adenine to normal mice stimulated blood and kidney levels of soluble tumor necrosis factor receptor 1 (sTNFR1) and kidney injury molecule-1 (KIM1), kidney hypertrophy, kidney mTOR activity, and kidney matrix production (Figures 4 G-K; Supplementary Figure 6).

### Endogenous adenine contributes to diabetic kidney disease in db/db mice

To determine whether endogenous adenine plays a role in progression of diabetic kidney disease, methylthio-DADMe-Immucillin-A (MTDIA) a small molecule specific inhibitor of methylthioadenosine phosphorylase (MTAP) was administered to db/db mice, a model of obese type 2 diabetes. MTAP converts methylthioadenosine to adenine and is responsible for approximately 80% of adenine production in mammalian cells (18). MTDIA was well tolerated and did not affect food intake, water intake, blood glucose levels or body weight (Supplementary Table 6). MTDIA significantly reduced kidney adenine in db/db mice (Figure 5A) but not other metabolites linked to progression of kidney disease (Supplementary Table 7) (19). MTDIA significantly reduced serum cystatin C, kidney hypertrophy, kidney KIM1, and partially reduced urine ACR, serum creatinine, urine KIM1, kidney matrix proteins and mTOR activity in db/db mice (Figure 5 B-I).

**Figure 5.**
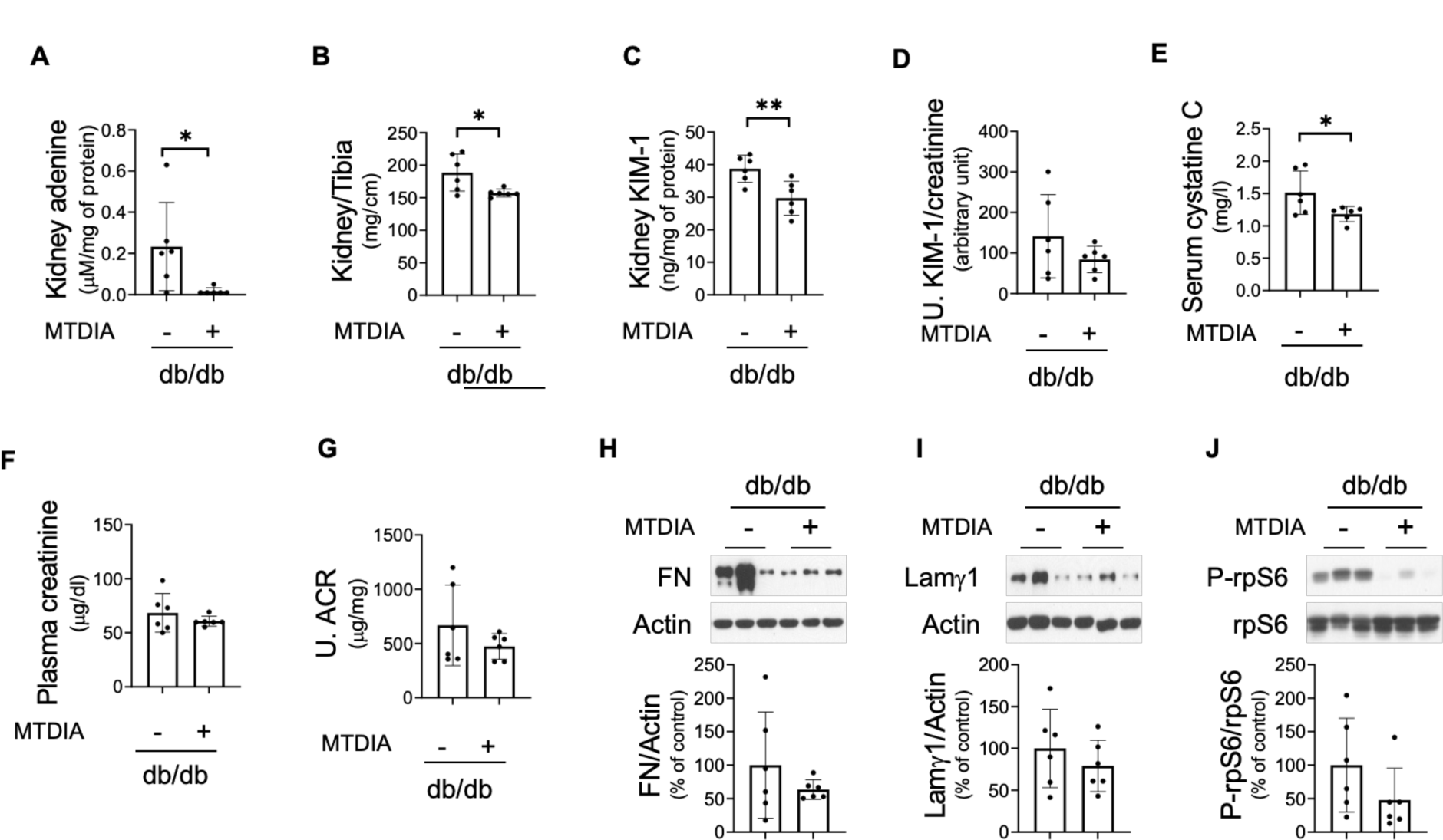
Methylthioadenosine phosphorylase (MTAP) inhibitor ameliorates kidney injury in db/db mice with type 2 diabetes. Methylthio-DADMe-Immucillin-A (MTDIA) significantly reduced kidney adenine levels (A), kidney hypertrophy (B), kidney KIM-1 levels (C) in diabetic mice. MTDIA significantly reduced diabetes-increased serum cystatin C (E), and partially reduced plasma creatinine (F), and albuminuria (G) in diabetic mice. Diabetes induced kidney matrix protein levels were partially reduced by MTDIA (H, I). Ribosomal S6 phosphorylation was partially reduced by MTDIA in the kidney of db/db mice (J) (n=6 per group, *p<0.05, **p<0.01, ***p<0.001).

## Discussion

The results from the present study demonstrate a role for endogenous adenine in kidney disease progression in the context of DKD. Urine levels of the AdCR identified patients with diabetes at high risk of kidney failure and all cause mortality at all levels of albuminuria in the CRIC study and verified in a cohort study from Singapore. The UAdCR can also identify patients who will develop ESKD even in the setting of normal or elevated GFR without macroalbuminuria across ethnicities. Spatial metabolomics localized adenine to regions of vascular, tubular, and glomerular pathology in patients with diabetes who have normoalbuminuria and GFR. Adenine appears to be in the causal pathway of kidney fibrosis as adenine was demonstrated to stimulate matrix molecules in proximal tubular cells via mTOR, was causative of kidney matrix production in mice and inhibiting adenine production was protective in diabetic mice.

Biomarkers in the causal pathway have not previously been identified for kidney disease progression in non-macroalbuminuric patients with diabetes. Microalbuminuria is clearly a risk factor for kidney disease progression, however as microalbuminuria can revert to normoalbuminuria (20) the dependence upon microalbuminuria alone may not provide reliable prognostication for event rates of GFR decline or kidney failure. Non-invasive omics approaches using plasma and urine have identified promising candidate biomarkers (21-23), however, demonstration of a contributory role of these biomarkers to the disease process has been difficult to establish (24). In the present study, integration of spatial metabolomics and single cell transcriptomics of human kidney biopsies converged on adenine and the mTOR pathway as highly relevant to DKD progression. The link with adenine and pathologic features was suggested by spatial metabolomics as adenine could be localized adjacent to atrophic tubules, in regions of arteriosclerotic blood vessels and glomerulosclerosis. The spatial localization implicated adenine to potentially be an endogenous pro-fibrotic factor.

Adenine is known to cause kidney pathology as an exogenous toxin in mouse (25) and rat models (14) of CKD, and possibly as an endogenous toxin in humans (26). The pathology of adenine-induced kidney disease includes glomerulosclerosis, tubular atrophy, interstitial fibrosis, and inflammatory cell infiltration (27, 28). The mechanism of adenine induced kidney disease has not been established although it has been postulated that conversion of adenine to 2-8 dihydroxyadenine (26) is a driver of CKD in patients with mutations of adenine phosphoribosyltransferase (APRT), the major enzyme that metabolizes adenine to AMP. However, CKD patients with APRT mutations are rare. Adenine itself is likely an endogenous tubular toxin based on the spatial metabolomic analysis and our finding that high urine adenine identifies patients at high risk of ESKD. Adenine exposure enhances tubular cell matrix production via the mTOR pathway and a prior study found that adenine is a potent stimulus for mTOR (17). Several published studies in mice and rats have also found that inhibiting mTOR protects against adenine-induced kidney disease (29-31). The mTOR pathway is likely relevant to human DKD as a recent study found stimulation of mTOR activity in kidney biopsies from patients with DKD (32) and our study with kidney biopsies from KPMP and CROCODILE demonstrate that a number of outputs of mTOR are elevated in DKD. This includes pathways involved in bioenergetics and pathways related to stimulation of extracellular matrix molecules. Further, adenine can increase the levels of KIM1 and sTNFR1 demonstrating that adenine is likely an initiator of downstream injury and inflammatory markers. Endogenous adenine production was blocked with a specific small molecule inhibitor of MTAP (MTDIA) and found to protect against diabetic renal hypertrophy, elevation of kidney KIM1 and was protective of decline in kidney function, as measured by serum cystatin C. It is possible that chronic MTAP inhibition with MTDIA could be developed as a safe therapeutic as a prior study found that MTDIA extended lifespan in mice with colon cancer and was provided for 294 days without evidence of toxicity (33). The role of adenine to accentuate mortality is not clear although it is possible that adenine could be directly toxic to vascular cells.

The measure of UAdCR was closely associated with DKD progression in the non-macroalbuminuric diabetic Pima Indian, CRIC, and SMART2D cohorts. As non-macroalbuminuric DKD leads to ESKD in many patients with CKD and diabetes (7, 8, 34), the new UAdCR biomarker could be of clinical value to identify those patients likely to progress. Further, the benefit of SGLT2 inhibitors may be due in part to reduce adenine levels as our study documented that short term use of empagliflozin significantly attenuated the UAdCR.

Strengths of our study included multiple analysis of several independent cohorts across different ages, ethnicities and stages of DKD. Additional strengths include application of spatial metabolomics and single cell transcriptomics to identify a pathway linking adenine to mTOR in human kidney disease pathology and progression. Limitations of our study was that the role of adenine was not demonstrated in type 1 DKD and other causes of CKD.

In conclusion, urine samples from independent well characterized cohorts of patients with diabetes identified the UAdCR as a robust predictor of ESKD and mortality independent of albuminuria and baseline eGFR, and spatial metabolomic and single cell transcriptomic studies from human kidney biopsies identified a potential role for endogenous adenine and the mTOR pathway in DKD. Studies in cells and mice identified a causative role for adenine and a small molecule therapeutic was found to block adenine production and was nephroprotective in a mouse model of type 2 diabetes. Our results thus demonstrate that endogenous adenine could contribute to progressive kidney disease in the context of type 2 diabetes.

## Methods

### Clinical Cohorts

Chronic Renal Insufficiency Cohort (CRIC): The parent CRIC Study recruited a racially diverse group aged 21 to 74 years, ∼50% with diabetes, with a broad range of kidney function (35). Informed consent was obtained from participants; protocols were approved by Scientific and Data Coordinating Center). The current study analyzed the urine sample at study entry (from baseline 24h urine samples) of 904 CRIC participants with diabetes and eGFR between 20-70 mL/min/1.73 m^2^ who had samples and outcomes data available. Singapore Study of Macro-Angiopathy and microvascular Reactivity in Type 2 Diabetes cohort (SMART2D): SMART2D is an ongoing prospective cohort study of Southeast Asian T2D participants recruited between 2011-2014 (36). Fasting spot urine samples were collected at baseline and stored at −80°C. To validate findings from the CRIC cohort, 309 participants with baseline eGFR 20-70 mL/min/1.73m^2^ and urine ACR < 300 µg/mg were evaluated. All participants gave written informed consent, and the study was approved by the Singapore National Healthcare Group. Pima Indians with early DKD were enrolled in a randomized clinical trial (37) (<u>ClinicalTrials.gov</u> number, <u>NCT00340678</u>). GFR was measured annually throughout the trial by the urinary clearance of iothalamate. Stored spot urine samples collected for two consecutive years were available from 54 participants and included for analysis. Additionally, urine samples were obtained under controlled euglycemic and hyperglycemic clamp conditions from a previously published clinical study in patients with type 1 diabetes (T1D) without macroalbuminuria (n=42) to evaluate the effects of Empagliflozin (Adjunctive-to-insulin and Renal Mechanistic (ATIRMA), NCT01392560) (38). Euglycemic clamp (4-6 mM glucose) conditions were maintained for approximately 4 hours prior to urine collection. The following day hyperglycemia (9-11 mM glucose) was maintained for 4 hours. Urine samples for adenine measurements were performed from samples obtained at the 4-hour time point following euglycemia or hyperglycemia before and after empagliflozin (25 mg/d) treatment for 8 weeks.

### Urine metabolomics (Zip-Chip Analysis)

Urine samples from Pima Indians, CRIC, SMART2D cohorts, and ATIRMA urines were all analyzed using ZipChip (908 Devices, Boston, MA) coupled with mass spectrometry (39). A rapid throughput urine adenine/creatinine assay was developed that showed excellent correlation with the gold standard assay using LC-MS/MS (Supplementary Figure 1). The reportable linear range for urine adenine assay was 100 nM to 100 uM, with a limit of detection at 10 nM and coefficient of variation (CV)<10% across the reportable linear range. Metabolite separation was achieved with a microfluidic chip which integrates capillary electrophoresis (CE) with nano-electrospray ionization through a ZipChip interface. Data acquisition was performed with Q-Exactive mass spectrometer (Thermo, San Jose, CA) and Thermo Scientific’s software Xcalibur-Quan Browser for data processing. Detailed procedures were previously published (39).

### Human kidney biopsies

Human kidney samples were obtained *via* the Kidney Precision Medicine Project (KPMP; The trial registration number from ClinicalTrials.gov is NCT04334707) and the Control of Renal Oxygen Consumption, Mitochondrial Dysfunction, and Insulin Resistance (CROCODILE) studies (40-42). The KPMP and CROCODILE studies were approved by the Institutional Review Board at Washington University, St. Louis, MO and the University of Colorado, respectively, and written consent was obtained from all patients. Samples were frozen in liquid nitrogen and stored at −80 °C until analysis. Snap frozen sample preparation and sectioning procedures for MALDI-MSI were published at dx.doi.org/10.17504/protocols.io.bcraiv2e.

### Animal studies

Zucker Diabetic Fatty (ZDF) rat kidney and urine samples were provided by Epigen, Inc. to verify that kidney spatial adenine correlated with the targeted urine adenine assay. C57Bl6J mice, db/m and db/db mice were obtained from Jackson Labs. C57Bl6J mice were administered adenine for 4 weeks in drinking water before sacrificing mice and harvesting tissues and blood samples after IACUC approval at UTHSA. db/m and db/db mice were administereted vehicle or methylthio-DADMe-Immucillin-A (MTDIA) MTAP inhibitor for a period of 8 weeks from week 10 to week 18. Albumin ELISA kit (Cat. #, E101 and E90-134, Bethyl Laboratories Inc.) and creatinine colorimetric kit (catalog ADI-907-030A, Enzo Life SciencesInc.) were used for the urinary ACR. Serum cystatin C was measured by Quantikine ELISA kit (Cat. #, MSCTC0, R&D systems). Plasma creatinine and metabolites in kidney tissue were measured by the ZipChip-mass spectrometry as previously described (19). Urine and kidney KIM-1 was measured by ELISA (Cat. #DY1817, R&D System).

### Mass Spectrometry Imaging (MSI) and Optical Imaging of Kidney Biopsies

A multimodal imaging approach was developed to investigate regional localization of metabolites in kidney sections. Bright-field (BF) and autofluorescence (AF) microscopy outlined glomeruli and tubules, and PAS-H staining revealed regions of pathology in serial sections. Matrix assisted laser desorption/ionization (MALDI)-MSI was performed with a Thermo Scientific Q Exactive HF-X hybrid quadrupole-Orbitrap mass spectrometer (Thermo Scientific) in combination with a novel elevated pressure MALDI/ESI interface (Spectroglyph LLC, Kennewick, WA) (43). Metabolite annotation was performed on METASPACE (44) and the optical image was uploaded to METASPACE and SCiLS Lab for visual overlay of metabolites with optical images to provide an assessment of metabolites associated with normal-appearing and pathologic features. Detailed procedures for MALDI-MSI are available at dx.doi.org/10.17504/protocols.io.bctfiwjn.

### Cell culture

Human kidney proximal tubular (HK-2) cells were purchased from American Type Culture Collection (Manassas, VA) and cultured as previously described (45). Murine kidney proximal tubular epithelial (MCT) cells were cultured as previously described (46). Cells were treated with 20 µM of adenine for the indicated time points with and without rapamycin (Fisher Scientific). Phosphorylation of S6 kinase, ribosomal protein S6 and expression of fibronectin and type 1 collagen α2 were analyzed by immunoblotting using antibodies against phosphor-Thr389-S6 kinase (Cat. # 9205, Cell Signaling Technology), phosphor-Ser240/244 ribosomal protein S6 (Cat. #, 2215, Cell Signaling Technology), ribosomal protein S6 (Cat. #, 2217, Cell Signaling Technology), MTAP (Cat. #, 62765, Cell Signaling Technology), fibronectin (Cat. #, ab2413, Abcam Plc), type 1 collagen α2 (Cat. #, 14695-1-AP, Proteintech Group Inc), and beta-actin (Cat. #, A2066, Millipore Sigma).

### Statistical Analysis

A composite kidney endpoint was defined as sustained kidney replacement therapy, progression to GFR or eGFR < 15 mL/min/1.73m^2^, or >50% GFR or eGFR decline from baseline level. All-cause mortality included death from any cause before reaching ESKD endpoint. Urine adenine was normalized to urine creatinine concentrations and log2 transformed. In the CRIC and SMART2D cohorts, the association of urine adenine levels (tertile) with clinical endpoints was studied by multivariable Cox proportional hazard regression models with adjustment for age, gender, ethnicity/race, body-mass index (BMI), hemoglobin A1c (HbA1c), mean arterial pressure (MAP), baseline eGFR, and urine ACR (natural log-transformed) as covariates. The group with a urine adenine/creatinine ratio in the lowest tertile was used as reference. Due to the limited number of cases in the Pima Indian cohort we only reported univariate Cox proportional hazards analysis for this cohort. To evaluate the pre-treatment and post-treatment effect of empagliflozin on urine adenine in the ATIRMA cohort, we performed a linear regression analysis for repeated measures.

### Bioinformatic and systems medicine analysis

Single cell transcriptomics and spatial metabolomics datasets generated from healthy kidney tissue, unaffected tissue in kidney nephrectomy and biopsy samples (KPMP and CROCODILE) were analyzed as recently described (40-42). The top pathway genes and proteins from the top 600 significant genes or proteins from proximal tubular cells were mapped onto pathways for subcellular processes using the MBCO ontology (47).

### Study Approval

For the CRIC study, Informed consent was obtained from participants; protocols were approved by IRBs and Scientific and Data Coordinating Center. All participants gave written informed consent, and the study was approved by the Singapore National Healthcare Group. The KPMP and CROCODILE studies were approved by the Institutional Review Board at Washington University, St. Louis, MO and the University of Colorado, respectively, and written consent was obtained from all patients.

## Author Contributions

The first 9 coauthors each made unique and critical contributions to this manuscript, and authorship order was determined after discussion among writing group members. KS, PB, SCL, JJL, RI, RGN, MK, and GZ designed the study. KS, PB, SCL, DC and RGN acquired funding for the study. GZ, JH, HJL, RM, PB, JJL, HL, LH, PB, LK, VSS acquired or generated the data. GZ, JH, HJL, JJL, HL, BC, LK and JG analyzed the data. KS, GZ, PB, MK, RGN and RI wrote the manuscript. KS, PB, MK, SCL, LN, JZ, VS, BK, SW, JH, KT, BK, TF, HF, IB, JS, HDL, JS, RM, EO, CA, TA, SCL, RGN, JG and RI provided scientific guidance and insights. All authors reviewed, edited, and approved the manuscript.

## Data Availability

All data produced in the present study are available upon reasonable request to the authors.

## Data Availability

All data produced in the present study are available upon reasonable request to the authors.

## Acknowledgments

G.Z, L.H., H.J.L, A.F, and K.S. receives salary and research support from NIH (UH3DK114920, 5U2CDK114886, RO1DK110541). L.N., J.Z., B.K. were supported by NIDDK 5R01DK110541. J.J.L. receives research support from Alexandra Health Fund (STAR grant 18203 and 20201). S.C.L. receives research support from Singapore National Medical Research Councile (MOH-000066, 0000714 and OFLCG/001/2017). P.B. receives salary and research support from NIDDK (R01 DK129211, R21 DK129720, K23 DK116720, UC DK114886, and P30 DK116073), JDRF (2-SRA-2019-845-S-B, 3-SRA-2017-424-M-B, 3-SRA-2022-1097-M-B), Boettcher Foundation, American Heart Association (20IPA35260142), Ludeman Family Center for Women’s Health Research at the University of Colorado, the Department of Pediatrics, Section of Endocrinology and Barbara Davis Center for Diabetes at University of Colorado School of Medicine. K.R.T. receives salary and research support from the NIDDK, NIMHD, NCATS, and NHLBI (R01MD014712, U2CDK114886, UL1TR002319, U54DK083912, U01DK100846, OT2HL161847, UM1AI109568) and the CDC (75D301-21-P-12254). R.G.N. was supported by the American Diabetes Association (Clinical Science Award 1-08-CR-42) and R.G.N. and H.C.L. were supported by the Intramural Research Program of NIDDK. J.H. and R.I. received salary support from U3CDK114886, R01GM137056, P01HL134605. Funding for the CRIC Study was obtained under a cooperative agreement from National Institute of Diabetes and Digestive and Kidney Diseases (U01DK060990, U01DK060984, U01DK061022, U01DK061021, U01DK061028, U01DK060980, U01DK060963, U01DK060902 and U24DK060990). In addition, this work was supported in part by: the Perelman School of Medicine at the University of Pennsylvania Clinical and Translational Science Award NIH/NCATS UL1TR000003, Johns Hopkins University UL1 TR-000424, University of Maryland GCRC M01 RR-16500, Clinical and Translational Science Collaborative of Cleveland, UL1TR000439 from the National Center for Advancing Translational Sciences (NCATS) component of the National Institutes of Health and NIH roadmap for Medical Research, Michigan Institute for Clinical and Health Research (MICHR) UL1TR000433, University of Illinois at Chicago CTSA UL1RR029879, Tulane COBRE for Clinical and Translational Research in Cardiometabolic Diseases P20 GM109036, Kaiser Permanente NIH/NCRR UCSF-CTSI UL1 RR-024131, Department of Internal Medicine, University of New Mexico School of Medicine Albuquerque, NM R01DK119199. We acknowledge technical help for animal studies from Richard Montellano, transcriptomics data organization from Fadhl AlAkwaa and Philip McCown, method development from Annapurna Pamreddy and data analysis by Rabiul Islam. D.C. has received operational funding for clinical trials from Boehringer Ingelheim-Lilly, Merck, Janssen, Sanofi, AstraZeneca, and Novo Nordisk.

## Appendix

The author affiliations are as follows: the Center for Precision Medicine, the University of Texas Health San Antonio, TX, USA (G.Z., A.F., L.H., H.J.L., M.A.V., C.A., J.G., K.S.), the Division of Nephrology, Department of Medicine, the University of Texas Health San Antonio, TX, USA (G.Z., H.J.L., K.S.), the Department of Pharmacological Sciences and Institute for Systems Biomedicine Icahn School of Medicine at Mount Sinai, New York, NY, USA (J.H., R.I.), the Chronic Kidney Disease Section, National Institute of Diabetes and Digestive and Kidney Diseases, Phoenix, AZ (H.C.L., R.G.N.), the Clinical Research Unit, Khoo Teck Puat Hospital, Singapore (J.J.L., S.C.L., J.G.), the Department of Population Health Sciences, University of Texas Health San Antonio, 7703 Floyd Curl Drive, San Antonio, TX, 78229, USA (R.F.), the SygnaMap, San Antonio, Texas, USA (L.H.), Audie L. Murphy Memorial VA Hospital, South Texas Veterans Health Care System, San Antonio, TX, USA (G.Z., H.J.L., K.S.), the Department of Pathology, University of Texas Health San Antonio, TX, USA (M.A.V.), the Department of Family Medicine and Public Health, Herbert Werthein School of Public Health, University of California-San Diego, La Jolla, California (L.N.), the University of California-San Diego Moores Cancer Center, University of California-San Diego, La Jolla, California (L.N., J.Z.), the Center for Molecular Medicine, Vienna, Austria (V.S.), the Department of Biostatistics, Fielding School of Public Health, University of California, Los Angeles, California (B.K.), the Division of Nephrology, Department of Medicine, Boston University, Boston, MA (S.W.), the Department of Medicine, University of Washington, Seattle, WA, USA and the Division of Nephrology, Department of Medicine, Kidney Research Institute, University of Washington, Seattle, Washington, USA (J.H., K.T., B.K., I.D.B.), the Division of Nephrology, Department of Medicine and Section of Endocrinology, Department of Pediatrics, University of Colorado Anschutz Medical Campus, Aurora, CO, USA (P.B.), the Institute of Molecular Systems Biology, ETH Zurich, 8093, Zurich, Switzerland (T.F.), the Center for Clinical Epidemiology and Biostistics, Perelman School of Medicine at the University of Pennsylvania and the Department of Biostatistics, Epidemiology, and Informatics, Perelman School of Medicine at the University of Pennsylvania (H.F.), the Epigen Biosciences, Inc., San Diego, California, USA (F.C.T.), the Cleveland Clinic, Cleveland, Ohio (J.S.), Department of Clinical Pharmacy and Pharmacology, University of Groningen, University Medical Center Groningen, Groningen, the Netherlands (H.L.H.), The George Institute for Global Health, Sydney, Australia (H.L.H.), the Department of Internal Medicine, University of Michigan, Ann Arbor, Michigan, USA (R.M., E.O., J.H., M.K.), the Environmental Molecular Sciences Laboratory, Pacific Northwest National Laboratory, Richland, WA, USA (C.A.), the Structural and Computational Biology Unit, European Molecular Biology Laboratory, Heidelberg, Germany (T.A.), the Diabetes Center, Admiralty Medical Center, Singapore and the Saw Swee Hock School of Public Health, National University of Singapore, Singapore and the Lee Kong Chian School of Medicine, Nayang Technological Univeristy, Singapore (S.C.L.)

---

**Supplementary Figure 1.**
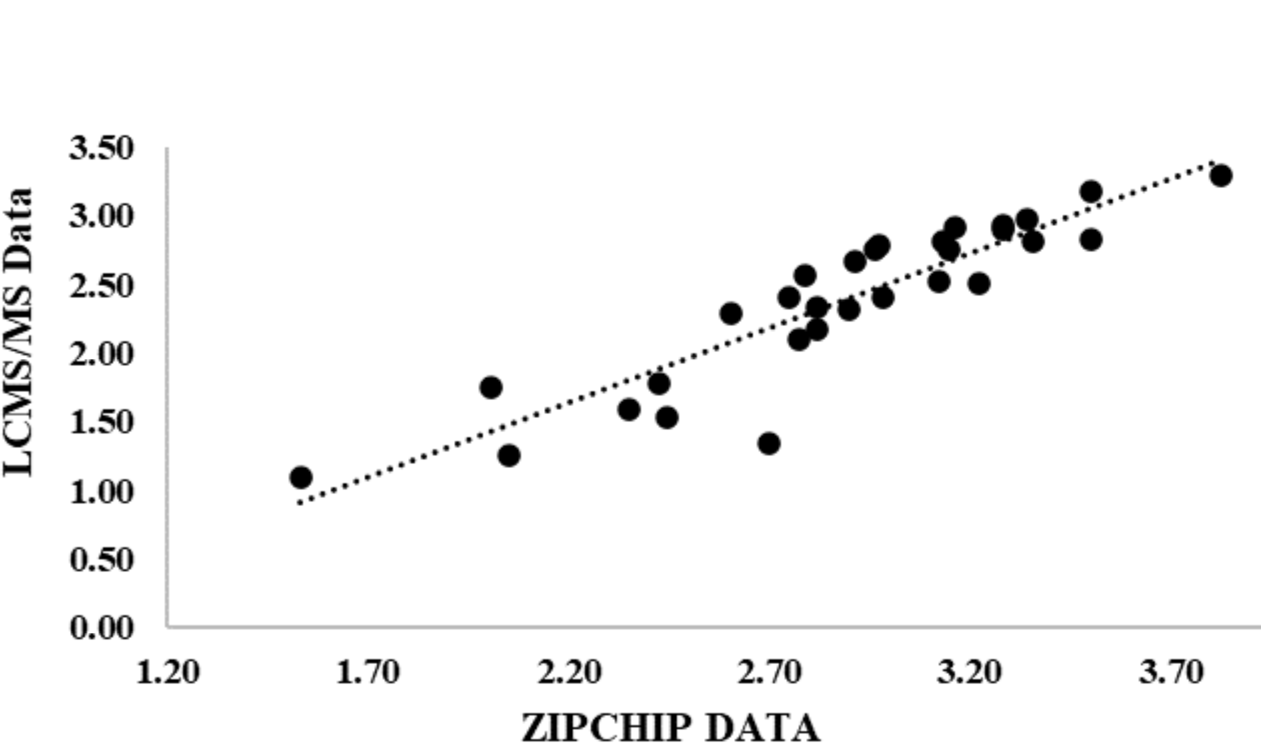
ZipChip urine adenine/creatinine assay correlates with LC-MS/MS. A ZipChip urine adenine/creatinine assay was developed and found to highly correlate with urine adenine/creatinine measured by LC-MS/MS. (n=23 samples, r=0.90, p<0.0001). Raw data was log-transformed and then normalized to urine creatinine. The unit for adenine concentration in the urine is nM/mM creatinine.

**Supplementary Figure 2.**
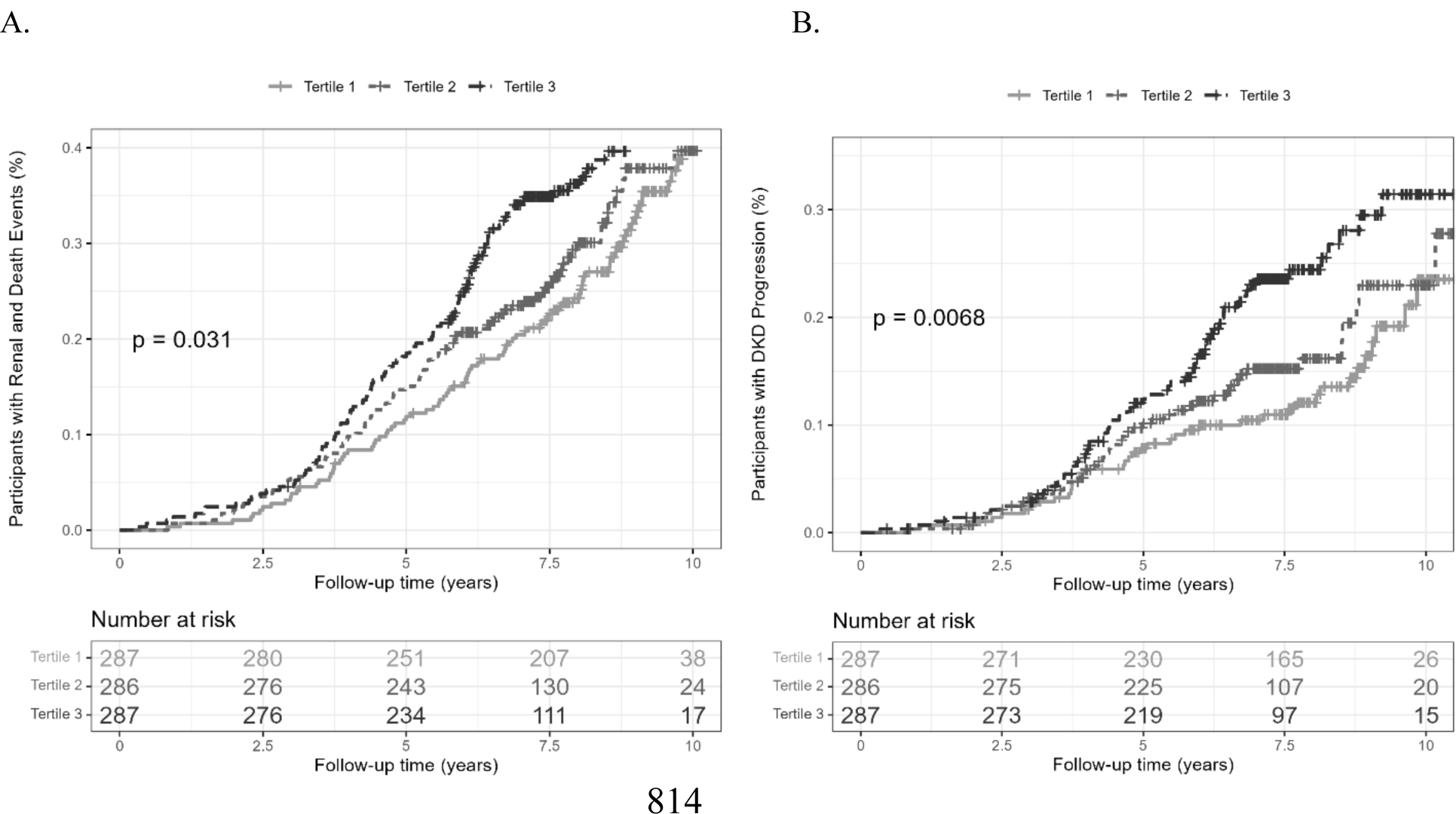
High urine adenine/creatinine ratio (UAdCR) levels identify patients with diabetes who are at high risk of ESKD and mortality (A) and ESKD (B) (combined CRIC and SMART2D). Participants with diabetes in the CRIC cohort and SMART2D cohorts had UAdCR levels measured within 1 year of enrollment and followed for 8-10 years. The top tertile had the highest risk of ESKD and all cause mortality (A) and ESKD alone (B).

**Supplementary Figure 3.**
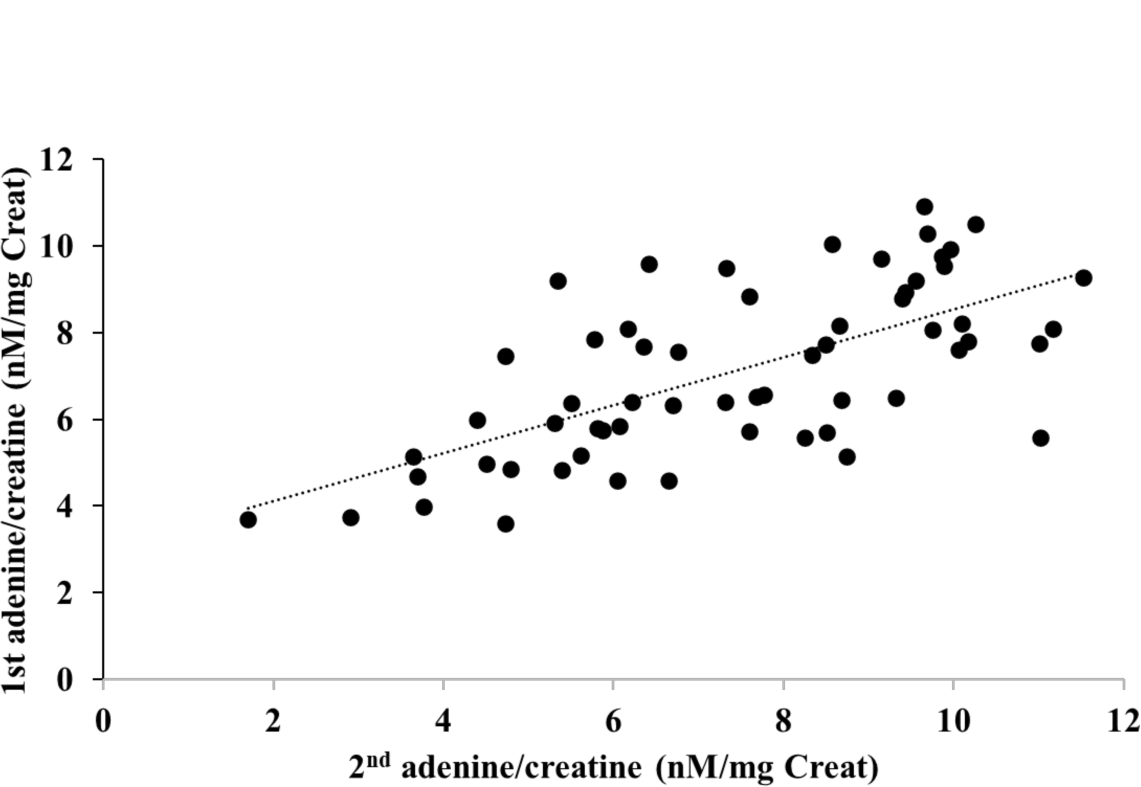
Urine adenine/creatinine ratio (UAdCR) levels are stable over time in Pima American Indians. Median time between measures of UAdCR is 12 months (IQR 11.8-12.5 months), R=0.665, p<0.0001, n=54 pairs).

**Supplementary Figure 4.**
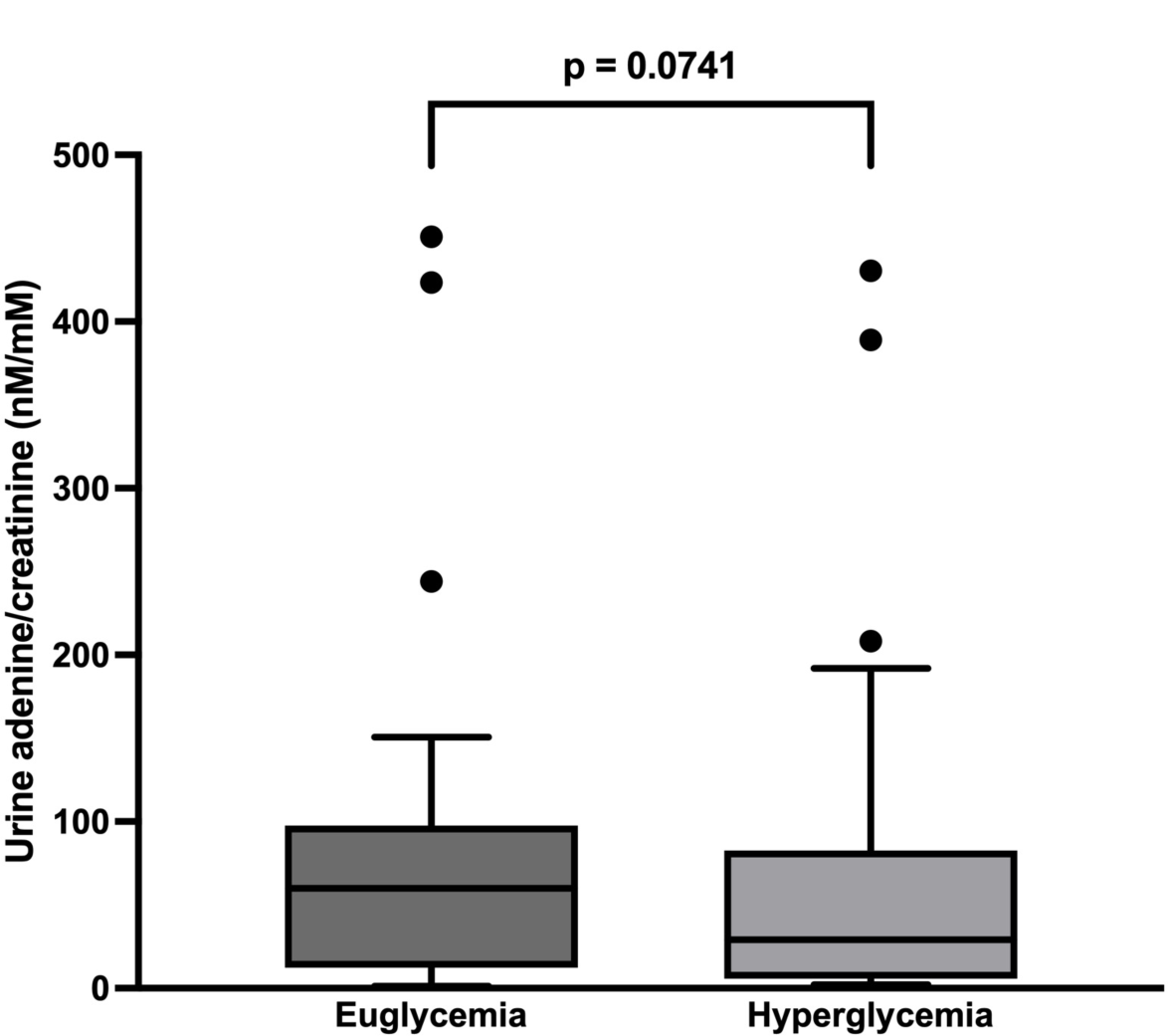
Hyperglycemia does not increase urine adenine/creatinine ratio (UAdCR) levels. Patients with T1 diabetes underwent euglycemic clamp or hyperglycemic clamp with UAdCR measured at end of clamp period (n=40 patients).

**Supplementary Figure 5.**
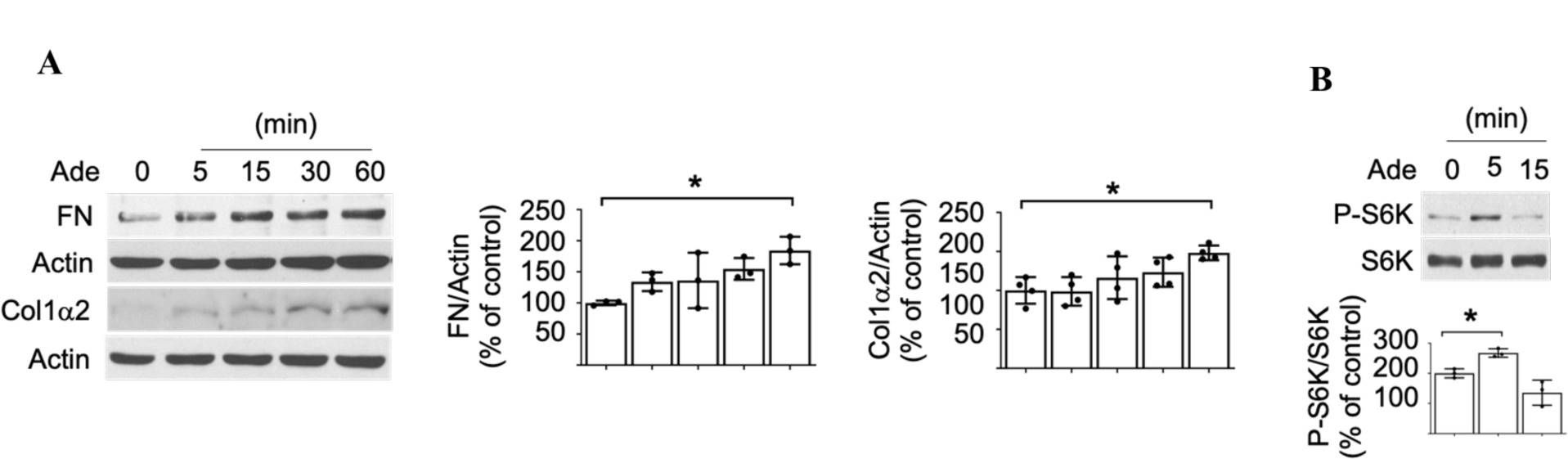
Adenine stimulates fibronectin, type I collagen and mTOR in human proximal tubular epithelial cells. Adenine administration to human kidney-2 (HK2) cells increases production of fibronectin and alpha2 chain of type I collagen (A) within 1h. Adenine stimulates mTOR activity within 5m of exposure as measured by phosphorylation of S6 kinase (B) (n=3 samples/group *p<0.05).

**Supplementary Figure 6.**
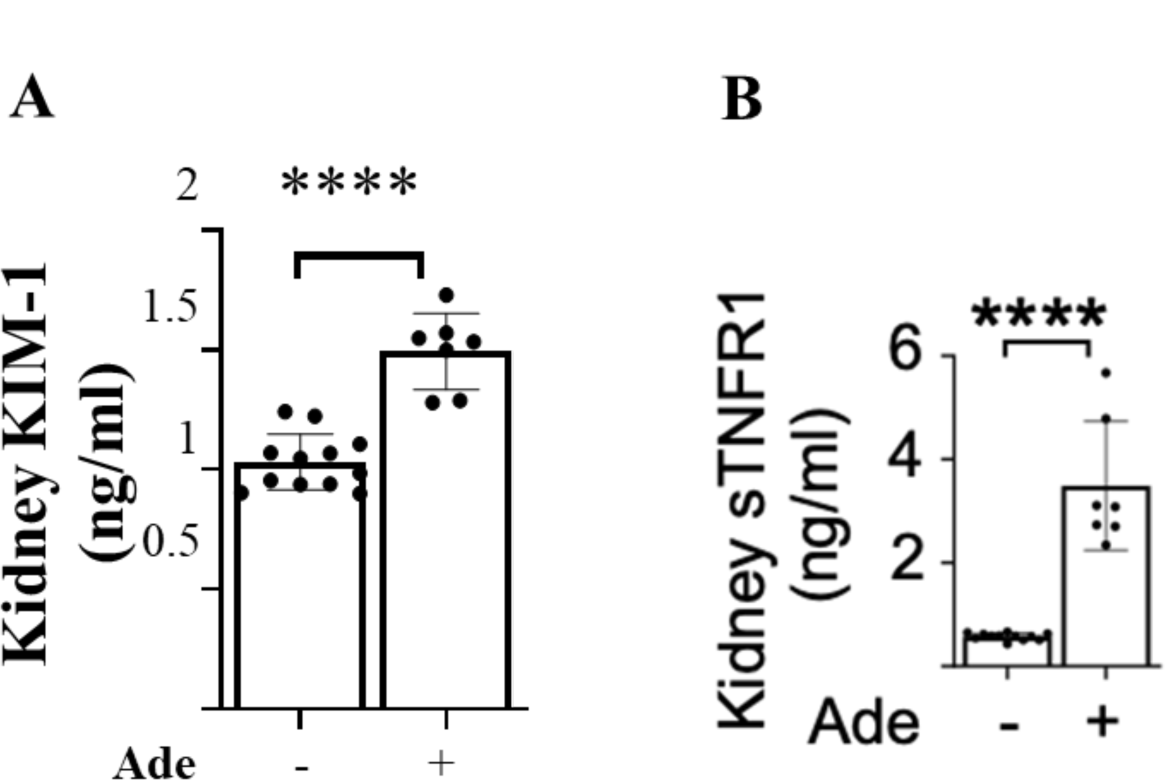
Adenine stimulates kidney KIM-1 and kidney sTNFR1 levels. Adenine administration to mice increases kidney levels of KIM-1 (A) and sTNFR1 (B). (n=12 in control group and n=7 in adenine treated group, ****p<0.001).

**Supplementary Table 1.** Hazard Ratios of ESKD or >50% decline in GFR incidence between tertiles of urine adenine/creatinine ratio (UAdCR) in Pima cohort without macroalbuminuria.

| Pima American Indians | N | Continuous |  |  | T2 vs T1 |  |  | T3 vs T1 |  |  |
| --- | --- | --- | --- | --- | --- | --- | --- | --- | --- | --- |
|  |  | HR | 95% CI | P value | HR | 95% CI | P value | HR | 95% CI | P value |
| No macroalbuminuria | 42 | 1.89 | 1.18-3.03 | 0.008 | 1.66 | 0.54-5.09 | 0.37 | 4.57 | 1.57-13.44 | 0.006 |

HR = hazard ratio; CI = confidence interval; No macroalbuminuria (ACR <300 mg/g). T1 = lowest tertile of UAdCR, T2=middle tertile of UAdCR and T3 = highest tertile of UAdCR. For the continuous measure of UAdCR, HR is per standard deviation in natural log-transformed UAdCR.

**Supplementary Table 2.** Association of baseline urine adenine/creatinine ratio (UAdCR) with risk for progression to ESKD in CRIC and SMART2D participants with non-macroalbuminuria and type 2 diabetes with 7 year follow up.

| Adenine/Creatinine ratio | CRIC cohort (N=551) |  | SMART2D cohort (N=309) |  |
| --- | --- | --- | --- | --- |
|  | HR (95% CI) | p value | HR (95% CI) | p value |
| 1-SD increment | 1.37 (1.08-1.73) | 0.011 | 1.70 (1.21-2.39) | 0.002 |
| Tertile 2 vs tertile 1 | 1.96 (1.06-3.60) | 0.031 | 0.91 (0.37-2.28) | 0.848 |
| Tertile 3 vs tertile 1 | 2.36 (1.26-4.39) | 0.006 | 2.39 (1.08-5.29) | 0.032 |

Multivariate Cox proportional hazard regression models were adjusted for baseline age, sex, ethnicity, body mass index, mean arterial pressure, hemoglobin A1c, eGFR and natural-log transformed urine albumin/creatinine ratio. UAdCR was modelled as both continuous variable (1-SD increment in log2-transformed adenine/creatinine ratio) and categorical variable (low tertile as reference). There were 7 subjects in CRIC with missing values for the clinical co-variates.

**Supplement Table 3.**
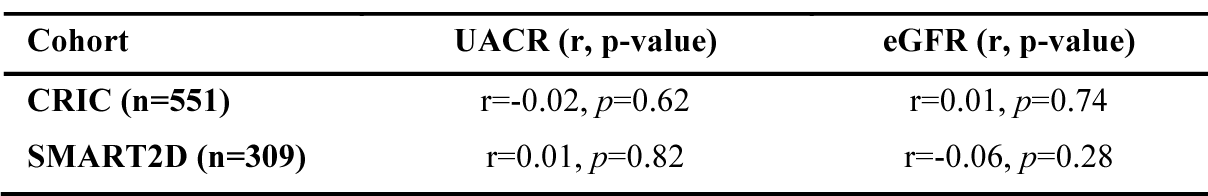
Correlations between urine adenine/creatinine ratio (UAdCR) and urine albumin/creatinine ratio (UACR) or the estimated glomerular filtration rate (eGFR) in the non-macro groups from CRIC and SMART2D.

**Supplementary Table 4.** Baseline characteristics of participants in ATIRMA, KPMP, and CROCODILE studies.

|  | ATIRMA (n=40) | KPMP (n=54) | CROCODILE (n=15) |
| --- | --- | --- | --- |
| Characteristics |  |  |  |
| Age, years | 24.3 ± 5.1 | (50.5 - 59.5) ± 12.7 | 24.1 ± 3.9 |
| Sex |  |  |  |
| Male, n (%) | 20 (50.0) | 22 (40) | 7 (46.7) |
| Weight, kg | 71.3 ± 12.6 | .. | 73.5 ± 12.7 |
| BMI, kg/m<sup>2</sup> | 24.1 ± 3.2 | .. | 25.2 ± 3.9 |
| HRT, n (%) | .. | 20 (36.4) | 5 (33.3) |
| T1D, n (%) | .. | 4 (7.3) | 8 (51.3) |
| T2D, n (%) | .. | 30 (56.4) | 2 (13.3) |
| HbA1C (%) | 7.8 ± 0.9 | .. | . |
| MAP, mmHg | 79.5 ± 7.4 | .. | . |
| eGFR*, mL/min/1.73m<sup>2</sup> | 149.8 ± 33.9 | (50.6 - 59.6) ± 23.6 | 148.8 ± 34.6 |
| Urine ACR (mg/g) | 10.35 ± 5.93 | .. | 7.8 ± 7.9 |

BMI: body-mass index, SBP: systolic blood pressure, DBP: diastolic blood pressure, MAP: mean arterial pressure, GFR: glomerular filtration rate, ERPF: estimated renal plasma flow, HbA1C: glycated hemoglobin A1c. (values expressed as Mean ± SD)

**Supplementary Table 5.** Correlation of urine adenine/creatinine ratio (UAdCR) with tissue levels of adenine.

| Spatial Adenine | Correlation with Urine Adenine/Creatinine with Kidney Adenine by MALDI-MSI |
| --- | --- |
| Glom Adenine | R = 0.64, p<0.001 |
| Non-Glom Adenine | R = 0.81, p<0.001 |
| Whole Slide Image Adenine | R = 0.73, p<0.001 |

Correlation of UAdCR measurements with tissue levels of adenine by spatial metabolomics of rat kidney samples (n=9).

**Supplementary Table 6.**
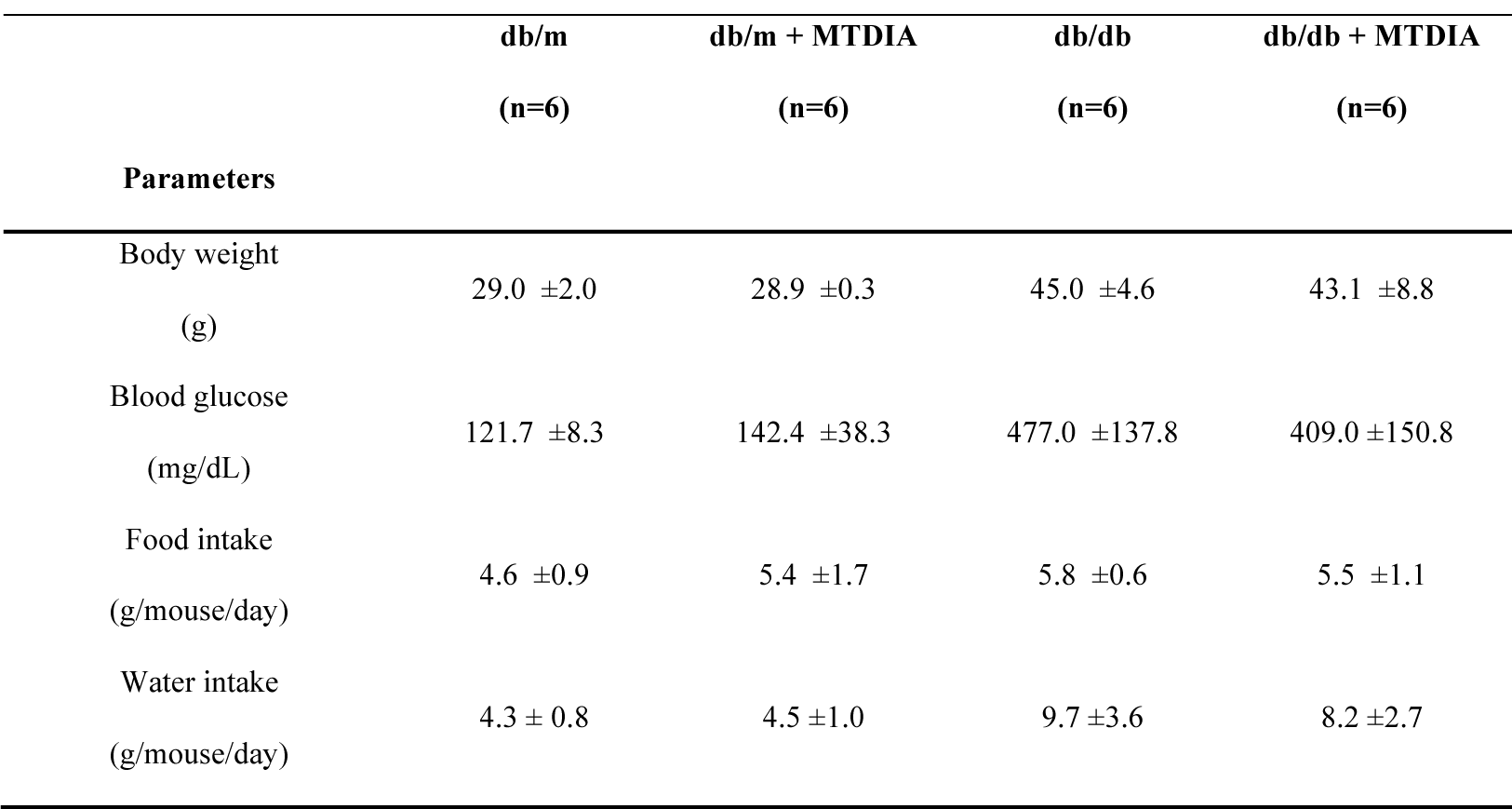
Clinical parameter in MTDIA treated db/db mice with type 2 diabets

**Supplementary Table 7.**
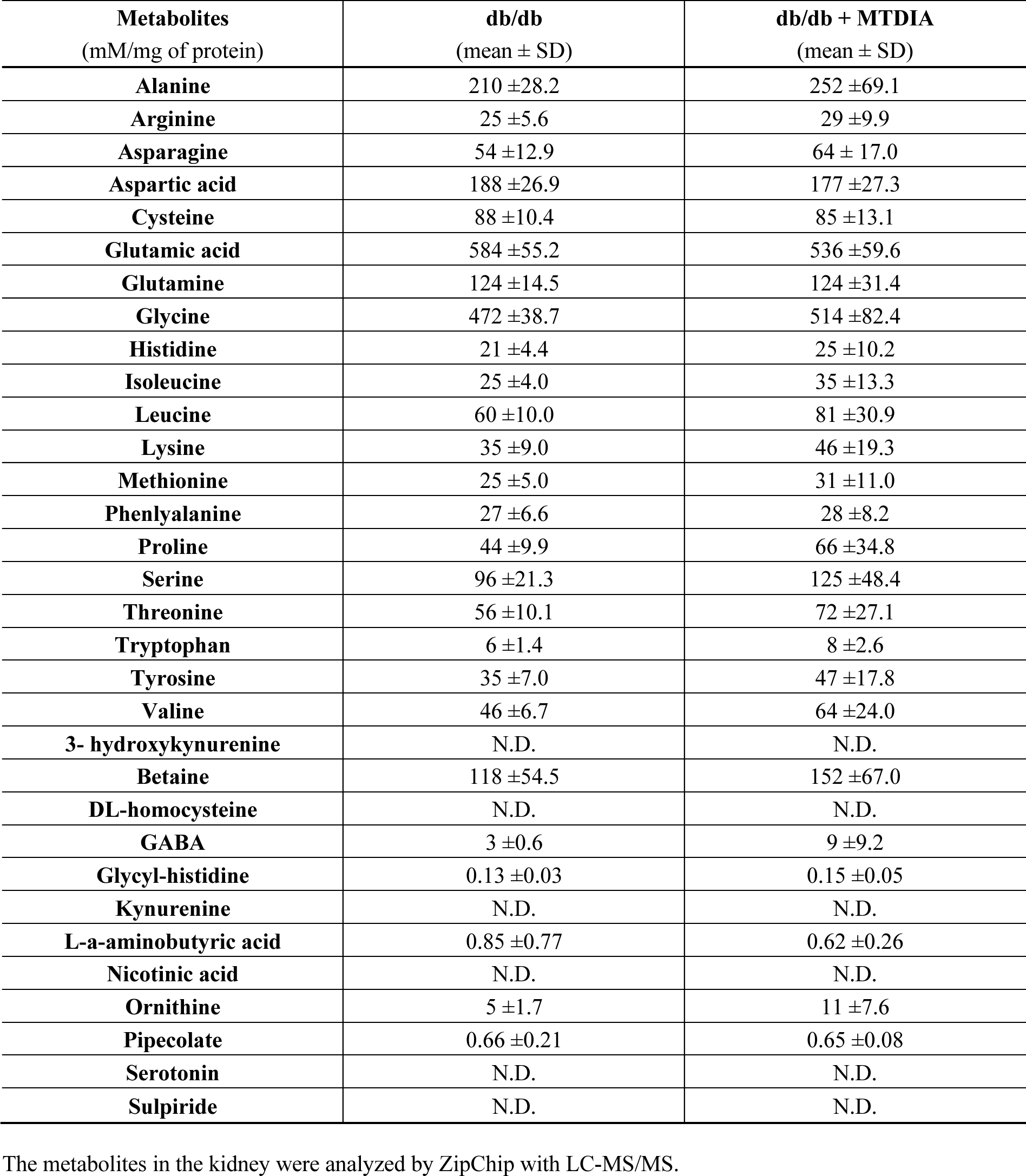
Kidney metabolites in MTDIA treated db/db mice with type 2 diabets

| **Metabolites**<br>(mM/mg of protein) | **db/db**<br>(mean ± SD) | **db/db + MTDIA**<br>(mean ± SD) |
| --- | --- | --- |
| **Alanine** | 210 ±28.2 | 252 ±69.1 |
| **Arginine** | 25 ±5.6 | 29 ±9.9 |
| **Asparagine** | 54 ±12.9 | 64 ± 17.0 |
| **Aspartic acid** | 188 ±26.9 | 177 ±27.3 |
| **Cysteine** | 88 ±10.4 | 85 ±13.1 |
| **Glutamic acid** | 584 ±55.2 | 536 ±59.6 |
| **Glutamine** | 124 ±14.5 | 124 ±31.4 |
| **Glycine** | 472 ±38.7 | 514 ±82.4 |
| **Histidine** | 21 ±4.4 | 25 ±10.2 |
| **Isoleucine** | 25 ±4.0 | 35 ±13.3 |
| **Leucine** | 60 ±10.0 | 81 ±30.9 |
| **Lysine** | 35 ±9.0 | 46 ±19.3 |
| **Methionine** | 25 ±5.0 | 31 ±11.0 |
| **Phenylalanine** | 27 ±6.6 | 28 ±8.2 |
| **Proline** | 44 ±9.9 | 66 ±34.8 |
| **Serine** | 96 ±21.3 | 125 ±48.4 |
| **Threonine** | 56 ±10.1 | 72 ±27.1 |
| **Tryptophan** | 6 ±1.4 | 8 ±2.6 |
| **Tyrosine** | 35 ±7.0 | 47 ±17.8 |
| **Valine** | 46 ±6.7 | 64 ±24.0 |
| **3- hydroxykynurenine** | N.D. | N.D. |
| **Betaine** | 118 ±54.5 | 152 ±67.0 |
| **DL-homocysteine** | N.D. | N.D. |
| **GABA** | 3 ±0.6 | 9 ±9.2 |
| **Glycyl-histidine** | 0.13 ±0.03 | 0.15 ±0.05 |
| **Kynurenine** | N.D. | N.D. |
| **L-a-aminobutyric acid** | 0.85 ±0.77 | 0.62 ±0.26 |
| **Nicotinic acid** | N.D. | N.D. |
| **Ornithine** | 5 ±1.7 | 11 ±7.6 |
| **Pipecolate** | 0.66 ±0.21 | 0.65 ±0.08 |
| **Serotonin** | N.D. | N.D. |
| **Sulpiride** | N.D. | N.D. |

The metabolites in the kidney were analyzed by ZipChip with LC-MS/MS.

## References

1. Nath KA. Tubulointerstitial changes as a major determinant in the progression of renal damage. Am J Kidney Dis. 1992;20(1):1–17.

2. Di Vincenzo A, Bettini S, Russo L, Mazzocut S, Mauer M, and Fioretto P. Renal structure in type 2 diabetes: facts and misconceptions. J Nephrol. 2020;33(5):901–7.

3. Mauer SM, Steffes MW, and Brown DM. The kidney in diabetes. Am J Med. 1981;70(3):603–12.

4. Caramori ML, Parks A, and Mauer M. Renal lesions predict progression of diabetic nephropathy in type 1 diabetes. J Am Soc Nephrol. 2013;24(7):1175–81.

5. Yamanouchi M, Furuichi K, Hoshino J, Ubara Y, and Wada T. Nonproteinuric diabetic kidney disease. Clinical and experimental nephrology. 2020;24(7):573–81.

6. Berhane AM, Weil EJ, Knowler WC, Nelson RG, and Hanson RL. Albuminuria and estimated glomerular filtration rate as predictors of diabetic end-stage renal disease and death. Clin J Am Soc Nephrol. 2011;6(10):2444–51.

7. Porrini E, Ruggenenti P, Mogensen CE, Barlovic DP, Praga M, Cruzado JM, et al. Non-proteinuric pathways in loss of renal function in patients with type 2 diabetes. Lancet Diabetes Endocrinol. 2015;3(5):382–91.

8. Pichaiwong W, Homsuwan W, and Leelahavanichkul A. The prevalence of normoalbuminuria and renal impairment in type 2 diabetes mellitus. Clin Nephrol. 2019;92(2):73–80.

9. Caramori ML, Fioretto P, and Mauer M. The need for early predictors of diabetic nephropathy risk: is albumin excretion rate sufficient? Diabetes. 2000;49(9):1399–408.

10. Forst T, Mathieu C, Giorgino F, Wheeler DC, Papanas N, Schmieder RE, et al. New strategies to improve clinical outcomes for diabetic kidney disease. BMC Med. 2022;20(1):337.

11. Tuttle KR, Agarwal R, Alpers CE, Bakris GL, Brosius FC, Kolkhof P, et al. Molecular mechanisms and therapeutic targets for diabetic kidney disease. Kidney Int. 2022;102(2):248–60.

12. Zhang J, Fuhrer T, Ye H, Kwan B, Montemayor D, Tumova J, et al. High-Throughput Metabolomics and Diabetic Kidney Disease Progression: Evidence from the Chronic Renal Insufficiency (CRIC) Study. Am J Nephrol. 2022:1–11.

13. Rahman A, Yamazaki D, Sufiun A, Kitada K, Hitomi H, Nakano D, et al. A novel approach to adenine-induced chronic kidney disease associated anemia in rodents. Plos One. 2018;13(2):e0192531.

14. Diwan V, Brown L, and Gobe GC. Adenine-induced chronic kidney disease in rats. Nephrology (Carlton). 2018;23(1):5–11.

15. Claramunt D, Gil-Pena H, Fuente R, Hernandez-Frias O, and Santos F. Animal models of pediatric chronic kidney disease. Is adenine intake an appropriate model? Nefrologia: publicacion oficial de la Sociedad Espanola Nefrologia. 2015;35(6):517–22.

16. Schiaffino S, Reggiani C, Akimoto T, and Blaauw B. Molecular Mechanisms of Skeletal Muscle Hypertrophy. J Neuromuscul Dis. 2021;8(2):169–83.

17. Hoxhaj G, Hughes-Hallett J, Timson RC, Ilagan E, Yuan M, Asara JM, et al. The mTORC1 Signaling Network Senses Changes in Cellular Purine Nucleotide Levels. Cell Rep. 2017;21(5):1331–46.

18. Lubin M, and Lubin A. Selective killing of tumors deficient in methylthioadenosine phosphorylase: a novel strategy. Plos One. 2009;4(5):e5735.

19. Zhang J, Fuhrer T, Ye H, Kwan B, Montemayor D, Tumova J, et al. High-Throughput Metabolomics and Diabetic Kidney Disease Progression: Evidence from the Chronic Renal Insufficiency (CRIC) Study. Am J Nephrol. 2022;53(2-3):215–25.

20. Perkins BA, Ficociello LH, Silva KH, Finkelstein DM, Warram JH, and Krolewski AS. Regression of microalbuminuria in type 1 diabetes. N Engl J Med. 2003;348(23):2285–93.

21. Niewczas MA, Pavkov ME, Skupien J, Smiles A, Md Dom ZI, Wilson JM, et al. A signature of circulating inflammatory proteins and development of end-stage renal disease in diabetes. Nat Med. 2019;25(5):805–13.

22. Tofte N, Lindhardt M, Adamova K, Bakker SJL, Beige J, Beulens JWJ, et al. Early detection of diabetic kidney disease by urinary proteomics and subsequent intervention with spironolactone to delay progression (PRIORITY): a prospective observational study and embedded randomised placebo-controlled trial. Lancet Diabetes Endocrinol. 2020;8(4):301–12.

23. Sharma K, Karl B, Mathew AV, Gangoiti JA, Wassel CL, Saito R, et al. Metabolomics reveals signature of mitochondrial dysfunction in diabetic kidney disease. J Am Soc Nephrol. 2013;24(11):1901–12.

24. Looker HC, Mauer M, and Nelson RG. Role of Kidney Biopsies for Biomarker Discovery in Diabetic Kidney Disease. Adv Chronic Kidney Dis. 2018;25(2):192–201.

25. de Frutos S, Luengo A, Garcia-Jerez A, Hatem-Vaquero M, Griera M, O’Valle F, et al. Chronic kidney disease induced by an adenine rich diet upregulates integrin linked kinase (ILK) and its depletion prevents the disease progression. Biochim Biophys Acta Mol Basis Dis. 2019;1865(6):1284–97.

26. George SA, Al-Rushaidan S, Francis I, Soonowala D, and Nampoory MRN. 2,8-Dihydroxyadenine Nephropathy Identified as Cause of End-Stage Renal Disease After Renal Transplant. Exp Clin Transplant. 2017;15(5):574–7.

27. Mo Y, Sun H, Zhang L, Geng W, Wang L, Zou C, et al. Microbiome-Metabolomics Analysis Reveals the Protection Mechanism of alpha-Ketoacid on Adenine-Induced Chronic Kidney Disease in Rats. Front Pharmacol. 2021;12:657827.

28. Saleh MA, Awad AM, Ibrahim TM, and Abu-Elsaad NM. Small-Dose Sunitinib Modulates p53, Bcl-2, STAT3, and ERK1/2 Pathways and Protects against Adenine-Induced Nephrotoxicity. Pharmaceuticals (Basel). 2020;13(11).

29. Wang J, Chai L, Lu Y, Lu H, Liu Y, and Zhang Y. Attenuation of mTOR Signaling Is the Major Response Element in the Rescue Pathway of Chronic Kidney Disease in Rats. Neuroimmunomodulation. 2020;27(1):9–18.

30. Nakano T, Watanabe H, Imafuku T, Tokumaru K, Fujita I, Arimura N, et al. Indoxyl Sulfate Contributes to mTORC1-Induced Renal Fibrosis via The OAT/NADPH Oxidase/ROS Pathway. Toxins (Basel). 2021;13(12).

31. Zhao Y, Zhao MM, Cai Y, Zheng MF, Sun WL, Zhang SY, et al. Mammalian target of rapamycin signaling inhibition ameliorates vascular calcification via Klotho upregulation. Kidney Int. 2015;88(4):711–21.

32. Schaub JA, AlAkwaa FM, McCown PJ, Naik AS, Nair V, Eddy S, et al. SGLT2 inhibitors mitigate kidney tubular metabolic and mTORC1 perturbations in youth-onset type 2 diabetes. J Clin Invest. 2023;133(5).

33. Firestone RS, Feng M, Basu I, Peregrina K, Augenlicht LH, and Schramm VL. Transition state analogue of MTAP extends lifespan of APC(Min/+) mice. Sci Rep. 2021;11(1):8844.

34. Liyanage P, Lekamwasam S, Weerarathna TP, and Srikantha D. Prevalence of normoalbuminuric renal insufficiency and associated clinical factors in adult onset diabetes. BMC Nephrol. 2018;19(1):200.

35. Yaffe K, Ackerson L, Tamura MK, Le Blanc P, Kusek JW, Sehgal AR, et al. Chronic Kidney Disease and Cognitive Function in Older Adults: Findings from the Chronic Renal Insufficiency Cohort Cognitive Study. Journal of the American Geriatrics Society. 2010;58(2):338–45.

36. Pek SL, Tavintharan S, Wang X, Lim SC, Woon K, Yeoh LY, et al. Elevation of a novel angiogenic factor, leucine-rich-alpha2-glycoprotein (LRG1), is associated with arterial stiffness, endothelial dysfunction, and peripheral arterial disease in patients with type 2 diabetes. J Clin Endocrinol Metab. 2015;100(4):1586–93.

37. Weil EJ, Fufaa G, Jones LI, Lovato T, Lemley KV, Hanson RL, et al. Effect of losartan on prevention and progression of early diabetic nephropathy in American Indians with type 2 diabetes. Diabetes. 2013;62(9):3224–31.

38. Škrtić M, Yang GK, Perkins BA, Soleymanlou N, Lytvyn Y, von Eynatten M, et al. Characterisation of glomerular haemodynamic responses to SGLT2 inhibition in patients with type 1 diabetes and renal hyperfiltration. Diabetologia. 2014;57(12):2599–602.

39. Liu H, Sridhar VS, Montemayor D, Lovblom LE, Lytvyn Y, Ye H, et al. Changes in plasma and urine metabolites associated with empagliflozin in patients with type 1 diabetes. Diabetes, obesity & metabolism. 2021;23(11):2466–75.

40. Lake BB, Chen S, Hoshi M, Plongthongkum N, Salamon D, Knoten A, et al. A single-nucleus RNA-sequencing pipeline to decipher the molecular anatomy and pathophysiology of human kidneys. Nat Commun. 2019;10(1):2832.

41. Menon R, Otto EA, Hoover P, Eddy S, Mariani L, Godfrey B, et al. Single cell transcriptomics identifies focal segmental glomerulosclerosis remission endothelial biomarker. JCI Insight. 2020;5(6).

42. Hansen J, Sealfon R, Menon R, Eadon MT, Lake BB, Steck B, et al. A reference tissue atlas for the human kidney. Science advances. 2022;8(23):eabn4965.

43. Belov ME, Ellis SR, Dilillo M, Paine MRL, Danielson WF, Anderson GA, et al. Design and Performance of a Novel Interface for Combined Matrix-Assisted Laser Desorption Ionization at Elevated Pressure and Electrospray Ionization with Orbitrap Mass Spectrometry. Anal Chem. 2017;89(14):7493–501.

44. Palmer A, Phapale P, Chernyavsky I, Lavigne R, Fay D, Tarasov A, et al. FDR-controlled metabolite annotation for high-resolution imaging mass spectrometry. Nature methods. 2017;14(1):57–60.

45. Yang L, Besschetnova TY, Brooks CR, Shah JV, and Bonventre JV. Epithelial cell cycle arrest in G2/M mediates kidney fibrosis after injury. Nat Med. 2010;16(5):535–43, 1p following 143.

46. Lee HJ, Lee DY, Mariappan MM, Feliers D, Ghosh-Choudhury G, Abboud HE, et al. Hydrogen sulfide inhibits high glucose-induced NADPH oxidase 4 expression and matrix increase by recruiting inducible nitric oxide synthase in kidney proximal tubular epithelial cells. J Biol Chem. 2017;292(14):5665–75.

47. Hansen J, Meretzky D, Woldesenbet S, Stolovitzky G, and Iyengar R. A flexible ontology for inference of emergent whole cell function from relationships between subcellular processes. Sci Rep. 2017;7(1):17689.

